# Exploiting convergent evolution to derive a pan-cancer cisplatin response gene expression signature

**DOI:** 10.1101/2021.11.10.21265799

**Authors:** Jessica A. Scarborough, Steven A. Eschrich, Javier Torres-Roca, Andrew Dhawan, Jacob G. Scott

## Abstract

Precision medicine offers remarkable potential for the treatment of cancer, but is largely focused on tumors that harbor actionable mutations. Gene expression signatures can expand the scope of precision medicine by predicting response to traditional (cytotoxic) chemotherapy agents without relying on changes in mutational status. We present a novel signature extraction method, inspired by the principle of convergent evolution, which states that tumors with disparate genetic backgrounds may evolve similar phenotypes independently. This evolutionary-informed method can be utilized to produce signatures predictive of response to over 200 chemotherapeutic drugs found in the Genomics of Drug Sensitivity in Cancer Database. Here, we demonstrate its use by extracting the Cisplatin Response Signature, CisSig, for use in predicting a common trait (sensitivity to cisplatin) across disparate tumor subtypes (epithelial-origin tumors). CisSig is predictive of cisplatin response within the cell lines and clinical trends in independent datasets of tumor samples. Finally, we demonstrate preliminary validation of CisSig for use in muscle-invasive cancer, predicting overall survival in patients who undergo cisplatin-containing chemotherapy. This novel methodology can be used to produce robust signatures for the prediction of traditional chemotherapeutic response, dramatically increasing the reach of personalized medicine in cancer.

**Translational Relevance:** Most precision medicine research focuses on using targeted drugs on patients with known driver mutations, yet the majority of patients don’t have actionable mutations. Using a novel signature extraction method, we produce the Cisplatin Response Signature (CisSig) to predict how well patients with epithelial-origin tumors will respond to cisplatin, a common cytotoxic chemotherapy. We show that expression of CisSig is correlated to clinical trends of cisplatin use in treatment guidelines using independent tumor databases. Then, we look at preliminary validation of CisSig for use in patients with muscle-invasive bladder cancer. Using two independent cohorts of pre-treatment tumor samples, we show that a CisSig-trained model is predictive of overall survival in patients who did receive cisplatin, but this signal is lost in patients who did not receive cisplatin–indicating that the model is predictive of therapeutic response, not simply prognosis.

## Introduction

Despite rich collections of cancer “-omic” data, precision medicine research has largely focused on producing therapies that target somatic mutations in previously documented driver genes. These therapies have produced some inspiring successes, extending the lives of patients with targetable mutations by months to years.^1–3^ However, the reach of genome-driven care is narrow and most patients without targetable mutations simply have not seen the benefits of personalized medicine. In fact, it was estimated that in 2020, just 7.04% of cancer patients in the United States could benefit from genome driven care.^4^ Even among the patients who do benefit from genome-driven care, the costs of these agents are high and the clinical responses are typically not durable, as tumors evolve in response to the targeted selection pressure, eventually becoming resistant to the drug.

Without an actionable mutation, patients often receive conventional cytotoxic chemotherapy. In these scenarios, there are significant opportunities for expanding the reach of precision medicine. For example, gene expression signatures can be used to predict response to these traditional chemotherapy agents without relying on changes in mutational status. Not only is gene expression a powerful measure of phenotype, it is readily translatable to a clinical setting, as patient tumors can undergo RNA-sequencing at relatively low cost and high scale. Defined as a set of genes (typically fewer than 100) whose expression covaries with a particular trait, certain gene expression signatures have already been incorporated into standard-of-care and clinical decision-making algorithms (e.g. OncotypeDx^5^, Mammaprint^6^). Additionally, signatures of radiosensitivity have been developed and have achieved level 1 evidentiary status for archival tissue.^7–10^

As seen in experimental and natural evolution, a variety of evolutionary trajectories can lead to the same phenotype.^11–14^ **Figure 1A** shows a canonical example of convergent evolution, where genomically disparate species (bats and birds) both evolved the same phenotype of flight independently of one another. Just as bats and birds are genetically closer to mice and reptiles, respectively, individual tumors may be genotypically similar to tumors with differing drug response phenotypes, **Figure 1B**. Under the selection pressure of a chemotherapeutic agent, tumors have many genotypes that may match to a given drug response phenotype, making a single genomic marker of drug sensitivity or resistance infeasible. In order to characterize chemotherapeutic response phenotype, our approach exploits the principles of convergent evolution by combining hundreds of cell lines from a variety of tissue types and extracting transcriptomic patterns of this phenotypic state.

**Figure 1.**
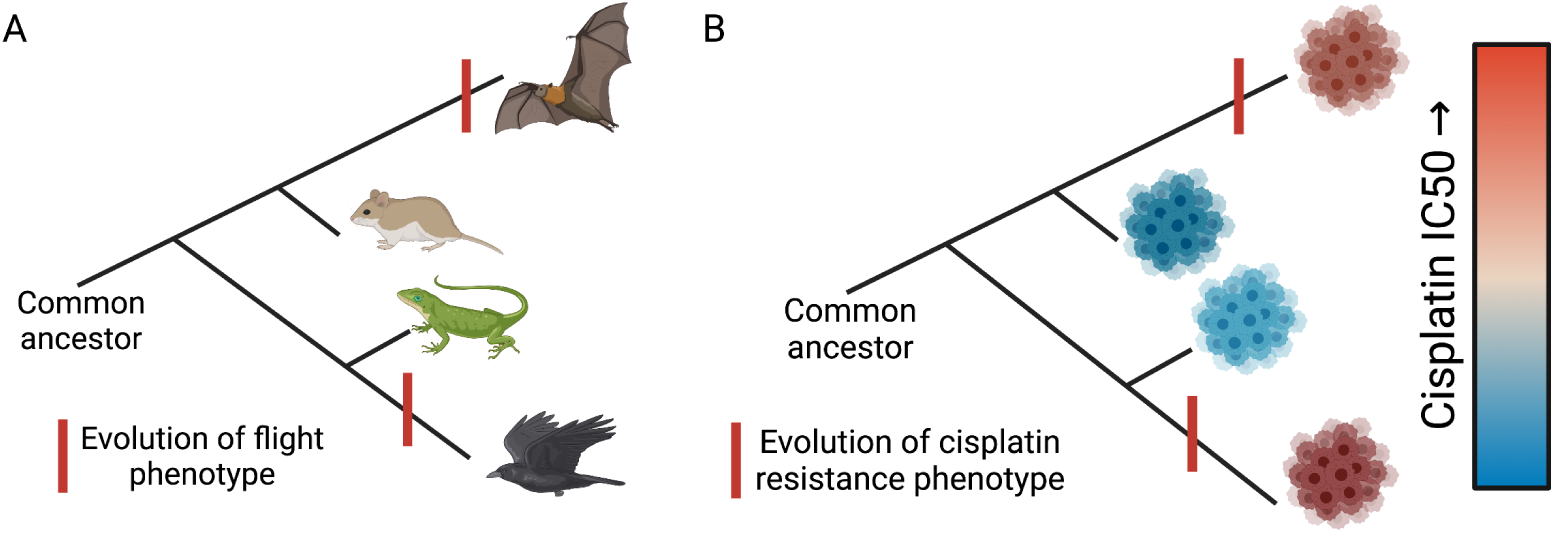
Visual representation of convergent evolution in animals and tumors. A. Birds and bats are genomically disparate, but both have individually evolved the ability to fly. B. Two tumors may evolve cisplatin resistance independently, despite being genomically distinct from one another.

While our novel method may be used to extract gene expression signatures for any quantitative or binary phenotype, here, we will demonstrate its utility with the extraction and validation of the Cisplatin Response Signature (CisSig), for use in predicting response to cisplatin in epithelial-origin tumors (carcinomas). Cisplatin is one of the most commonly used chemotherapy agents, given to variety of cancer subtypes including bladder, head and neck, gynecological, and many more disease sites. Given its widespread use, it comes as no surprise that prior work has assessed the utility of mutational and transcriptomic signatures in predicting response to this drug; yet, to the best knowledge of these authors, none of these advancements have been translated into routine clinical care.^15–19^ Furthermore, in contrast to our pan-epithelial strategy, most previously published cisplatin response signatures or biomarkers are intended for application in a single disease site.

This work employs a seed gene approach, as in Buffa et al., where previously identified hypoxia-regulated genes became seeds in a co-expression network, and highly connected genes formed a hypoxia metagene (gene signature).^20^ By extracting genes that are highly co-expressed with biologically significant genes, Buffa et al. produced a robust hypoxia gene signature which was prognostic, even in multivariate analysis and across multiple tissue types.^21, 22^

Our method derives these seed genes using differential gene expression analysis, comparing cisplatin-sensitive and -resistant cell lines from the Genomics of Drug Sensitivity in Cancer (GDSC) database. Of note, this empirical approach to gene extraction is distinct from the majority of signature extraction methods, which rely on genes with a known role in drug response or cancer development. The seed genes are trimmed based on co-expression in epithelial-based tumor samples from The Cancer Genome Atlas (TCGA) ensuring that the final signature contains genes that tend to be expressed together in both cell lines and clinical samples. We then show that our final signature is highly predictive of drug response within GDSC cell lines, and we establish that signature expression is congruent with use of cisplatin in standard of care guidelines between disease sites. And finally, we provide an example of how CisSig may be translated for use in a single disease site, muscle-invasive bladder cancer (MIBC), predicting which patients will not benefit from cisplatin-containing chemotherapy.

## Results

### Convergent evolution informs Cisplatin Response Signature (CisSig) derivation

CisSig was derived using 429 epithelial-based cancer cell lines in the GDSC Database, each characterized for gene expression and drug response (see **Figure 2A**). The distribution of disease sites for these cell lines may be found in **Supplementary Table 2**. GDSC gene expression consists of RMA normalized microarray data, details discussed in Methods. This database reports both half-maximal inhibitory concentration (IC50) and area under the drug response curve (AUC) as measures of drug response. A Spearman correlation between these two metrics demonstrated reasonable concordance (*ρ* = 0.84, p *<<* 0.001) in measuring cisplatin response for our cell lines of interest (**Supplementary Fig. 1**). We therefore moved forward with IC50 as the metric of drug response, as it is a more commonly reported measure.

**Figure 2.**
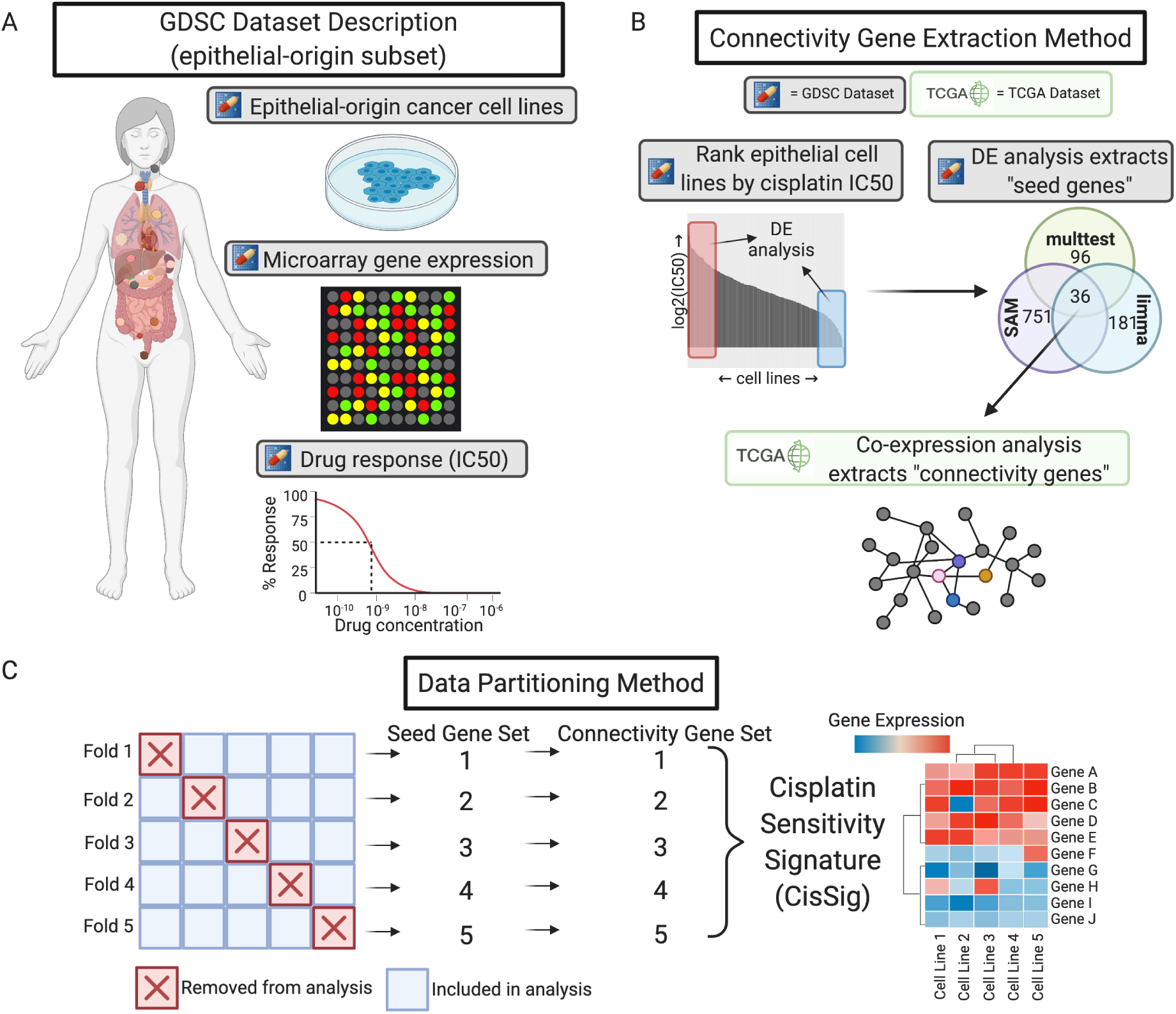
Schematic representation of CisSig derivation. **A.** Description of the epithelial-origin subset of the Genomics of Drug Discovery in Cancer (GDSC) dataset (denoted with the pill icon in future figures). These data include 429 epithelial-based cancer cell lines, with drug response measurements to over 200 drugs and gene expression characterization via microarray. **B.** Pipeline for extracting connectivity seeds. First, differential gene expression analysis between the top and bottom 20% of cisplatin responders found genes with significantly increased expression in a state of cisplatin sensitivity. These differentially expressed genes became “seed genes” in a co-expression network built using gene expression from clinical samples of epithelial-based tumors in The Cancer Genome Atlas (TCGA). Seed genes that were highly co-expressed with each other were denoted as “connectivity genes.” **C.** Schematic of data partitioning, where GDSC epithelial-based cancer cell lines from **A.** are split into 5 folds. Each fold underwent the pipeline in **B.** Genes found in at least 3 of the 5 connectivity gene sets were included in the final signature, CisSig.

Each of these folds was analyzed with a pipeline of differential gene expression and co-expression analysis, visually depicted in **Figure 2B** and discussed below. This pipeline was performed across five partitions of the data, each with a different 20% of the cell lines removed (each containing 343 or 344 cell lines), illustrated in **Figure 2C**. The method utilized multiple partitions of the data in order to find genes that are consistent between folds, reducing the chance for outlier cell lines to influence the results.

With no pre-filtering of genes, differential gene expression (DE) analysis using limma,^23^ SAM,^24^ and multtest^25^ methods was performed between the top and bottom 20% of responders (i.e. cell lines with the highest and lowest 20% of IC50 values). The distribution of disease sites found in each comparison group (resistant and sensitive) for each fold may be found in **Supplementary Tables 2-6**. More details on parameters and version numbers for each DE method can be found in the Methods section. For each fold, the genes found to be over-expressed in a cisplatin-sensitive state by all three DE methods were termed the “seed genes,” resulting in 5 sets of seed genes, as depicted in **Figure 2C**. Using only intersecting genes between the three methods is done with the goal of increasing stringency by reducing overall false discovery rate. Results of the DE analysis for each fold are summarized in **Supplementary Table 7**, and lists of differentially expressed genes from each method, for each fold can be found in Supplementary Data.

A co-expression network was built for each set of seed genes, as described in Methods and visually represented in the bottom panel of **Figure 2B**. These networks were built using The Cancer Genome Atlas (TCGA) RNA-Seq expression data from epithelial-based tumor samples, comparing the expression of each seed gene and all other genes in the dataset. Seed genes that were highly co-expressed with each other are extracted from each fold, termed “connectivity seeds.” Here, we bring in gene expression from tumor samples (not cell lines) to ensure that only genes that are expressed together in both cell lines and tumor samples are included in the final signature. The final gene signature, CisSig, contains any gene found in at least 3 of the 5 sets of connectivity seeds, and the genes included in the signature are listed in **Table 1**.

**Table 1.** Genes included in CisSig. These genes all appear in at least 3 of the 5 sets of connectivity seeds.

| HGNC Gene Symbol | Gene Name |
| --- | --- |
| *ADAT2* | Adenosine Deaminase tRNA Specific 2 |
| *ATP1B3* | ATPase Na<sup>+</sup>/K<sup>+</sup> transporting subunit beta 3 |
| *CDIN1* | CDAN1 interacting nuclease 1 |
| *C1QBP* | Complement C1q binding protein |
| *CDC7* | Cell division cycle 7 |
| *CDCA7* | Cell division cycle associated 7 |
| *FKBP14* | FKBP prolyl isomerase 14 |
| *KRT5* | Keratin 5 |
| *LRRC8C* | Leucine rich repeat containing 8 VRAC subunit C |
| *LY6K* | Lymphocyte antigen 6 family member K |
| *MMP10* | Matrix metalloproteinase 10 |
| *NPM3* | Nucleophosmin 3 |
| *PSAT1* | Phosphoserine aminotransferase 1 |
| *RIOK1* | RIO kinase 1 |
| *SLFN11* | Schlafen family member 11 |
| *STOML2* | Stomatin like 2 |
| *USP31* | Ubiquitin specific peptidase 31 |
| *WDR3* | WD repeat domain 3 |
| *ZNF750* | Zinc finger protein 750 |

Using the ‘sigQC’ package in R, we analyzed a suite of quality control metrics to assess the robustness of CisSig in a clinical sample (TCGA) dataset.^26, 27^ The signature is compared to the 5 sets of seed genes originally extracted from GDSC prior to being refined by co-expression analysis. These results are visualized in a radar plot in **Supplementary Figure 2**. CisSig demonstrates greater intra-signature correlation, increased correlation between mean and median, and decreased skewness within RNA-expression from TCGA samples of epithelial origin. Other metrics of interest include the coefficient of variance and the proportion (*σ*) of signature genes found in the top 10%, 25% or 50% of variable genes. These metrics can be used to assess the variability of signature genes within a dataset, where it is ideal to have signature genes that vary more than the background noise. Here, CisSig performs similarly to the unfiltered differential gene expression results. Finally, the these metrics are summarized into a score, also displayed in **Supplemental Figure 2**, where CisSig outperformed all sets of seed genes.

### Increased CisSig expression predicts cisplatin sensitivity within GDSC dataset

**Figure 3A** demonstrates the expression of CisSig genes in cisplatin-sensitive and -resistant GDSC cell lines (top and bottom IC50) quintiles. From this, we see that signature expression tends to be higher (more red) in sensitive, rather than resistant, cell lines. Next, a “CisSig score,” the median normalized expression of the 19 CisSig genes, is calculated for the same sensitive and resistant cell lines. The distribution of CisSig score and IC50 among all cell lines can be found in **Supplementary Figure 3**.

**Figure 3.**
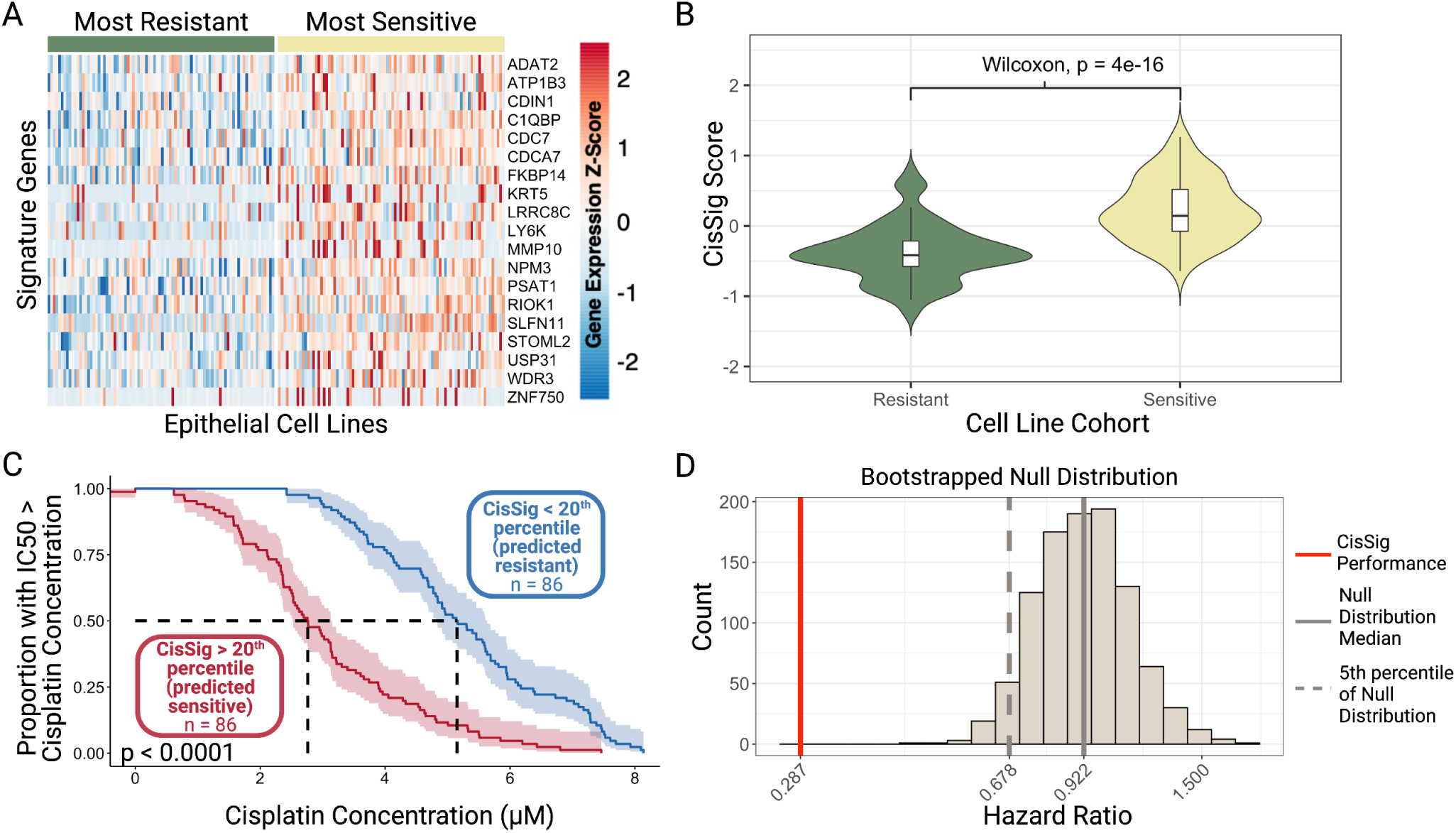
Visualization of CisSig expression within GDSC Dataset. **A.** An unclustered heatmap showing gene expression of the CisSig genes (rows) in cell lines (columns) from the top and bottom quintiles of cisplatin IC50. Color of the heatmap represents the Z-score of gene expression, normalized to each gene. Cell lines denoted as sensitive (right, yellow bar) tend to display higher expression of CisSig genes than cell lines denoted as resistant (left, green bar). Z-scores above 2.5 are denoted as 2.5, and Z-scores below -2.5 are denoted as -2.5. **B.** Violin plots comparing the distribution of CisSig scores between the cell lines in the highest and lowest quintile of cisplatin IC50. A Wilcoxon Rank Sum Test found that the median CisSig scores between these two cohorts was significantly different (p < 0.001). **C.** Comparison of the distribution cisplatin IC50 between cell lines in the highest and lowest quintile of CisSig score. Y-axis represents the proportion of the cohort with a cisplatin IC50 greater than the cisplatin concentration on the X-axis. A log-rank test between the two cohorts demonstrates significantly different drug response between the two cohorts (p < 0.0001). **D.** Null distribution of hazard ratio using 1000 random gene signatures with the same length as CisSig and the model described in **C.** CisSig’s performance is compared to the 95% confidence interval of the null distribution, where each signature’s performance (CisSig and nulls) is represented by the hazard ratio between two cohorts separated by the signature score.

**Figure 3B** shows that sensitive cell lines tend to have higher CisSig scores than resistant cell lines. This is expected, given that the seed genes were initially extracted as genes with increased expression in a cisplatin-sensitive state in the GDSC dataset.

**Figure 3C** compares the distribution of IC50 between cohorts of GDSC cell lines in this top and bottom quintile of CisSig score. We are terming this plot a “Cell Line Persistence Curve,” which resembles a Kaplan-Meier survival curve, but uses IC50 in place of survival time for cell lines. Here, we assume that a cell line does not “survive” when the concentration of cisplatin is greater than it’s IC50. For example, at 50% “survival” on the y-axis, the median IC50 of the high CisSig cohort is 2.76 *µM* (left, vertical dashed line), while the median IC50 of the low CisSig cohort is 5.15 *µM* (right, vertical dashed line). In other words, cell lines predicted to be resistant (low CisSig) tend to have greater IC50 values and cell lines predicted to be sensitive (high CisSig) tend to have lower IC50 values.

As demonstrated by Venet et al, many published gene signatures do not perform significantly better when predicting survival outcomes than random gene signatures of the same length^28^. Given the large sample size of cell lines, simply testing for statistical significance may not be stringent enough. Therefore, we compared the performance of CisSig’s Cell Line Persistence Curve (hazard ratio) to the performance of a null distribution. This null distribution was created using 1000 random gene signatures with the same length as CisSig, assessing the hazard ratio between each signature’s Cell Line Persistence Curve. In **Figure 3D**, we see that CisSig drastically outperforms the top 95% of this null distribution.

### CisSig Score is not related to status of common DNA damage response genes

We compared the expression of CisSig to a variety of genes that are commonly associated with response to DNA damage, such as the application of a cytotoxic chemotherapy like cisplatin. The genes we examined include, but are not limited to, BRCA1/2, PTEN, RAD51C/D, and ATM. In Supplementary Figure 4, we show a heatmap of mutation status for 16 genes across all epithelial-based cell lines in the GDSC dataset. Each column in the heatmap represents a GDSC cell line, ordered from high CisSig Score to low CisSig Score). The presence of a mutation in any of the genes does not appear to be more or less common as CisSig Score increases, indicating that CisSig may represent biomarker information that is orthogonal to mutation status of DNA damage response genes.

### CisSig outperforms the null distributions of drug response prediction models in the GDSC dataset

In **Figure 3C-D**, we demonstrated a novel method to show the stark difference in IC50 distribution for GDSC cell lines with high and low CisSig scores, but it is also important to assess CisSig’s predictive power using more traditional methods. To that aim, we built a variety of prediction models using CisSig to predict IC50 as a continuous or binary outcome in epithelial-based GDSC cell lines, described in **Table 2**. We chose to evaluate the efficacy of using a summary score (CisSig score) in addition to individual gene expression in order to show the value of more “basic” statistical models (e.g. simple linear regression) for producing an easier to interpret model while also gauging the power of using individual CisSig genes in accurately predicting drug response (e.g. random forest). When utilizing expression of each gene individually as the input for our models, we chose a penalized form of regression to prevent overfitting. Finally, for each method selected, we chose to build two models, one with all epithelial-based cell lines and one with only epithelial-based cell lines with high or low signature expression (based on CisSig score quintiles). In doing so, we can gauge whether more extreme expression of CisSig is related to improved drug response prediction accuracy.

**Table 2.** Model details and validation results for the prediction of cisplatin response using CisSig in GDSC dataset.

| Input | Output | Method | Included Data | Metric | Value |
| --- | --- | --- | --- | --- | --- |
| CisSig Score | IC50 (continuous) | Simple Linear Regression | All | Corr. Coef. | 0.51 |
| CisSig Score | IC50 (continuous) | Simple Linear Regression | Quintiles | Corr. Coef. | 0.74 |
| All gene expression | IC50 (continuous) | Elastic Net Linear Regression | All | Corr. Coef. | 0.63 |
| All gene expression | IC50 (continuous) | Elastic Net Linear Regression | Quintiles | Corr. Coef. | 0.79 |
| All gene expression | IC50 (continuous) | L1 Linear Regression | All | Corr. Coef. | 0.63 |
| All gene expression | IC50 (continuous) | L1 Linear Regression | Quintiles | Corr. Coef. | 0.79 |
| All gene expression | IC50 (continuous) | L2 Linear Regression | All | Corr. Coef. | 0.63 |
| All gene expression | IC50 (continuous) | L2 Linear Regression | Quintiles | Corr. Coef. | 0.81 |
| All gene expression | IC50 (binary) | Simple Logistic Regression | All | AUC | 0.79 |
| All gene expression | IC50 (binary) | Simple Logistic Regression | Quintiles | AUC | 0.90 |
| All gene expression | IC50 (binary) | Elastic Net Logistic Regression | All | AUC | 0.82 |
| All gene expression | IC50 (binary) | Elastic Net Logistic Regression | Quintiles | AUC | 0.94 |
| All gene expression | IC50 (binary) | L1 Logistic Regression | All | AUC | 0.82 |
| All gene expression | IC50 (binary) | L1 Logistic Regression | Quintiles | AUC | 0.94 |
| All gene expression | IC50 (binary) | L2 Logistic Regression | All | AUC | 0.81 |
| All gene expression | IC50 (binary) | L2 Logistic Regression | Quintiles | AUC | 0.95 |
| All gene expression | IC50 (binary) | SVM (linear kernel) | All | AUC | 0.82 |
| All gene expression | IC50 (binary) | SVM (linear kernel) | Quintiles | AUC | 0.93 |
| All gene expression | IC50 (binary) | SVM (polynomial kernel) | All | AUC | 0.78 |
| All gene expression | IC50 (binary) | SVM (polynomial kernel) | Quintiles | AUC | 0.94 |
| All gene expression | IC50 (binary) | Random Forest | All | AUC | 0.81 |
| All gene expression | IC50 (binary) | Random Forest | Quintiles | AUC | 0.91 |

In short, simple linear regression models used CisSig score to predict a cell line’s IC50 as a continuous variable, while elastic net, L1-, and L2-penalized linear regression models used expression of all CisSig genes to predict a cell line’s IC50 as a continuous variable. For these linear regression models, performance was compared using the Spearman correlation coefficient (*ρ*) between the predicted and actual IC50 value for the cell lines withheld from a given fold’s training dataset. The best correlation coefficient between the five folds is chosen to represent each model, shown in **Table 2**. Simple logistic regression models used CisSig score to predict a cell line’s IC50 as a binary outcome (above or below the median), while elastic net-, L1-, and L2-penalized logistic regression, support vector machine (with linear and polynomial kernels), and random forest models were built to use expression of each CisSig gene to predict IC50 as a binary outcome. We used area under the receiver operating characteristic (ROC) curve (AUC) to represent each classification model’s performance, again choosing the best of five folds to represent the model in **Table 2**.

All models demonstrate improved performance when trained and tested on only cell lines with the highest and lowest signature scores. Additionally, the penalized regression models outperform the simple regression models when comparing the same cell line data inputs. It is expected that including CisSig genes as individual variables would improve performance in comparison to CisSig score, but it is noteworthy that something as simple as median normalized expression of all CisSig genes (also known as the CisSig score) could predict IC50 with the performance shown here.

**Figure 4** shows the performance of CisSig for each of the modeling methods described in **Table 2**. In **Figures 4A-B**, we demonstrate how each of the violin plots in **Figures 4C-D** were built. For example, in **Figure 4A**, we assess a linear regression model with CisSig score from all epithelial-based GDSC cell lines as the input and IC50 as the continuous outcome. Each model is built with five-fold cross validation, and performance is measured by comparing the predicted and actual IC50 of the testing set using a Spearman correlation. The best performance of the five-folds is used to represent CisSig’s performance, shown in **Figure 4A**. Next, a null distribution, shown in **Figure 4B**, is produced using 1000 random gene signatures with the same length as CisSig and the same modeling method. Again, the best performance of the five-folds is used to represent each null signature’s performance, and CisSig is compared to the null distribution.

**Figure 4.**
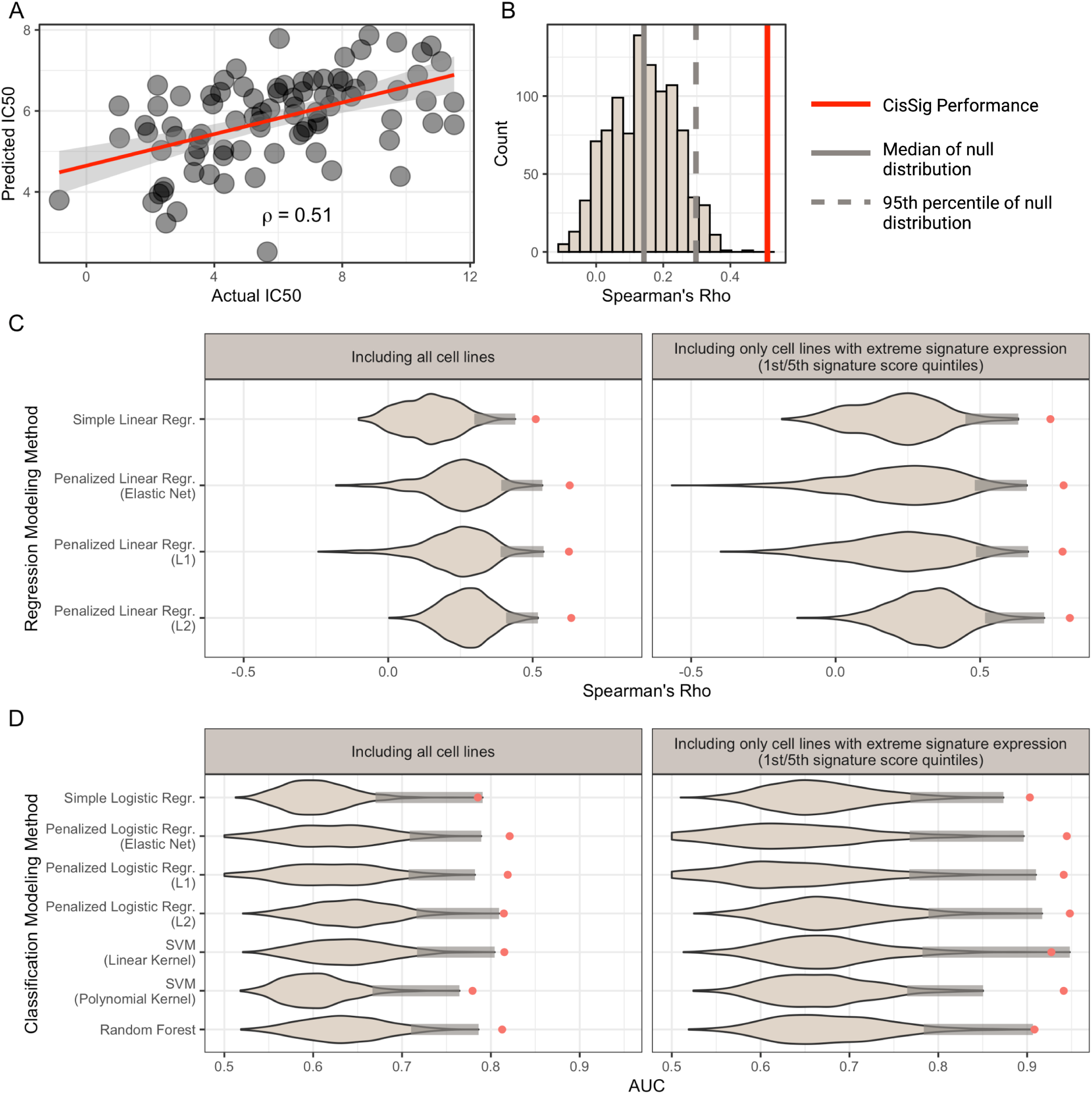
CisSig predicts IC50 using a variety of modeling techniques in the GDSC dataset. **A.** Scatterplot of the actual vs. predicted IC50 using CisSig score to predict IC50 with linear regression. Plot shows the best performing fold (measured by Spearman’s rho) from 5-fold cross validation. **B.** Null distribution of the performance metric from **A.** (Spearman’s rho), built using 1000 random gene signatures to predict IC50 as described in **A.** As with CisSig, the metric of the best performing fold is used to represent each null signature. The median of the null distribution and the cutoff for the 95th percentile of the null distribution are represented by the solid and dashed gray line, respectively. CisSig’s performance, red solid line, outperforms at least 95% of the null distribution. **C.** Violin plots containing the null distribution of performance metrics for 11 modeling methods. Each distribution was created as discussed in **A-B**, where CisSig’s performance is compared to the performance of 1000 random gene signatures of the same length. For each violin, a shaded gray bar represents the top 5% of each null distribution and CisSig’s performance is shown with a red dot. The modeling methods, including input and output, are described in Table 2.

We repeated the modeling described in **Figures 4A-B** for 10 additional modeling methods and the two versions of the dataset (one including all cell lines and another including only cell lines in the top and bottom quintile of signature expression). In **Figures 4C-D**, we show that CisSig outperforms these null distributions for each of the 11 modeling methods using both versions of the dataset, often outperforming the null distribution altogether. Finally, **Supplementary Figures 5-15** presents CisSig’s performance in each of the cross validation folds and show a detailed histogram of each model’s null distribution.

**Figure 5.**
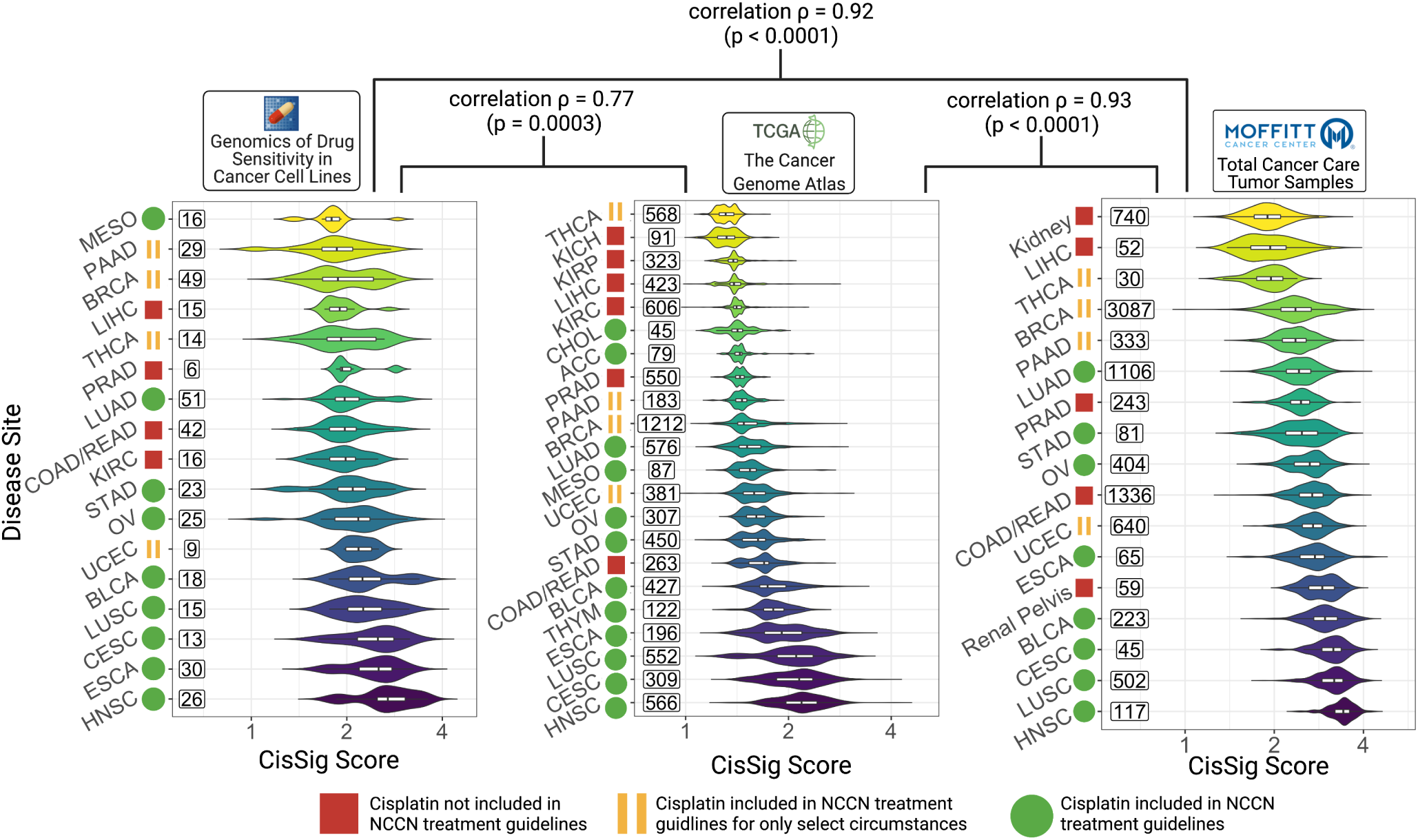
Cancer subtypes with greater CisSig expression tend to have cisplatin included in standard of care guidelines. Cancer subtypes are ranked by median CisSig Score in three data sets, GDSC (left), TCGA (middle), and TCC (right). The color of each violin plot represents the rank of the cancer subtype. The ranks of intersecting subtypes between each dataset are compared with Spearman’s rank correlation, reported with correlation *ρ* and p-value. Rank correlation *ρ* between GDSC and TCGA and GDSC and TCC datasets is 0.77 (p = 0.0003) and 0.902 (p « 0.0001). Rank correlation *ρ* between TCGA and TCC datasets is 0.93 (p « 0.0001). Violin plots display the distribution of CisSig scores for each cancer subtype. Within each violin, a boxplot denotes median signature score for each subtype (middle horizontal line) and 25th/75th percentile for signature scores (box edges). Numbers to the left of each violin plot represent sample size included in each cancer subtype.

A wide variety of modeling methods is included in this analysis in order to demonstrate that although no one method is predictably superior to another, CisSig shows strong predictive power when utilizing any of them. Additionally, models that include only cell lines with more extreme signature expression consistently have improved performance compared to the same modeling method that includes all cell lines. This intimates that more extreme CisSig expression can more accurately predict a cell line’s response to cisplatin.

### Ranking cancer subtypes by CisSig expression is concordant with observed clinical trends

In addition to demonstrating strong utility in predicting the drug response of epithelial-based cell lines, CisSig’s expression was examined across disease sites in external clinical samples. Using three large datasets, we assessed how expression of CisSig relates to cisplatin use across epithelial-based cancer disease sites. CisSig score was calculated for all samples (cell lines or clinical tumor samples) in GDSC, TCGA, and Total Cancer Care (TCC) databases. In order to visualize these scores on a log-transformed axis, signature score was linearly scaled, such that the lowest score became exactly 1.

In **Figure 5**, disease sites were ranked by the median signature score for the cohort in GDSC (left), TCGA (middle), and TCC (right) datasets. Furthermore, each disease site is labeled as utilizing cisplatin in NCCN treatment guidelines (green circle), using cisplatin in very select circumstances (yellow bars), or not having cisplatin included in NCCN treatment guidelines (red square). In all datasets, we see that disease sites with higher CisSig scores tend to have cisplatin included in treatment guidelines, while those with lower scores tend to not have cisplatin included in treatment guidelines.

Finally, disease site rank was compared between datasets using Spearman’s correlation. There is a strong correlation between the rank of shared disease sites of all three datasets. Between GDSC and TCGA, Spearman’s *ρ* is 0.77 (p < 0.001). Between GDSC and TCC, Spearman’s *ρ* is 0.92 (p < 0.001). And between TCGA and TCC, Spearman’s *ρ* is 0.93 (p < 0.001). This high degree of concordance between datasets signifies that CisSig displays consistent expression between a variety of data sources (including between microarray and RNA-seq methods).

### CisSig is predictive of survival in muscle-invasive bladder cancer (MIBC) patients who received cisplatin containing chemotherapy

We trained and tested a Cox proportional hazards (PH) survival model using CisSig genes with two publicly available datasets, described in in Table 3. Both sets of tumor samples used the same platform for gene expression profiling. Within Dataset A, we performed univariate survival analysis with each of the CisSig genes using only samples that received cisplatin-containing neo-adjuvant chemotherapy. Genes with a a strong relationship between increased expression and improved survival (as seen in GDSC cell lines) were selected to be included in multivariate analysis; for additional details, see Methods.

**Table 3.** Description of clinical datasets used for training and testing of CisSig-informed survival model. Treatment refers to neoadjuvant MVAC chemotherapy, which is a regimen that includes methotrexate, vinblastine, doxorubicin, and cisplatin. Gene expression profiling in both datasets was performed using the Illumina HumanHT-12 WG-DASL V4.0 R2 expression beadchip platform.

| Name | GSE Accession No. | Disease Site | *n* with treatment | *n* without treatment |
| --- | --- | --- | --- | --- |
| Dataset A | GSE48276 | Bladder | 16 | 37 |
| Dataset B | GSE70691 | Bladder | 22 | 0 |

As shown in Figure 6A, this multivariate analysis used Dataset A samples that received cisplatin-containing treatment, producing a trained Cox PH model. We tested this model using samples from Dataset B, which also received cisplatin-containing chemotherapy and the samples from Dataset A that did not receive cisplatin-containing chemotherapy. Figures 6B-C show that samples predicted to be “high risk” have significantly worse survival than patients predicted to be “low risk.” Figure 6B uses an arbitrary cutoff (median) to separate the cohorts, while Figure 6C uses the optimal cutoff to separate the groups. Similarly, Figures 6D-E show significant separation between “high,” “medium,” and “low risk” cohorts with worst to best survival outcomes, respectively. Again, Figure 6D uses an arbitrary cutoff (tertiles) to separate the cohorts, while Figure 6E uses the optimal two cutpoints for each cohort. Finally, Figures 6F-G show that the signal is lost when testing our model with either binary or tertile cohorts in patients from Dataset A who did not receive cisplatin-containing chemotherapy. The reverse of these analyses, where the model is trained with Dataset B’s patients who did receive cisplatin-containing chemotherapy, then is tested using Dataset A’s patients who both did and did not receive cisplatin-containing chemotherapy shows similar results, shown in Supplementary Figure 16. For both models, the coefficients and their standard errors can be found in Supplementary Tables 9 and 10.

**Figure 6.**
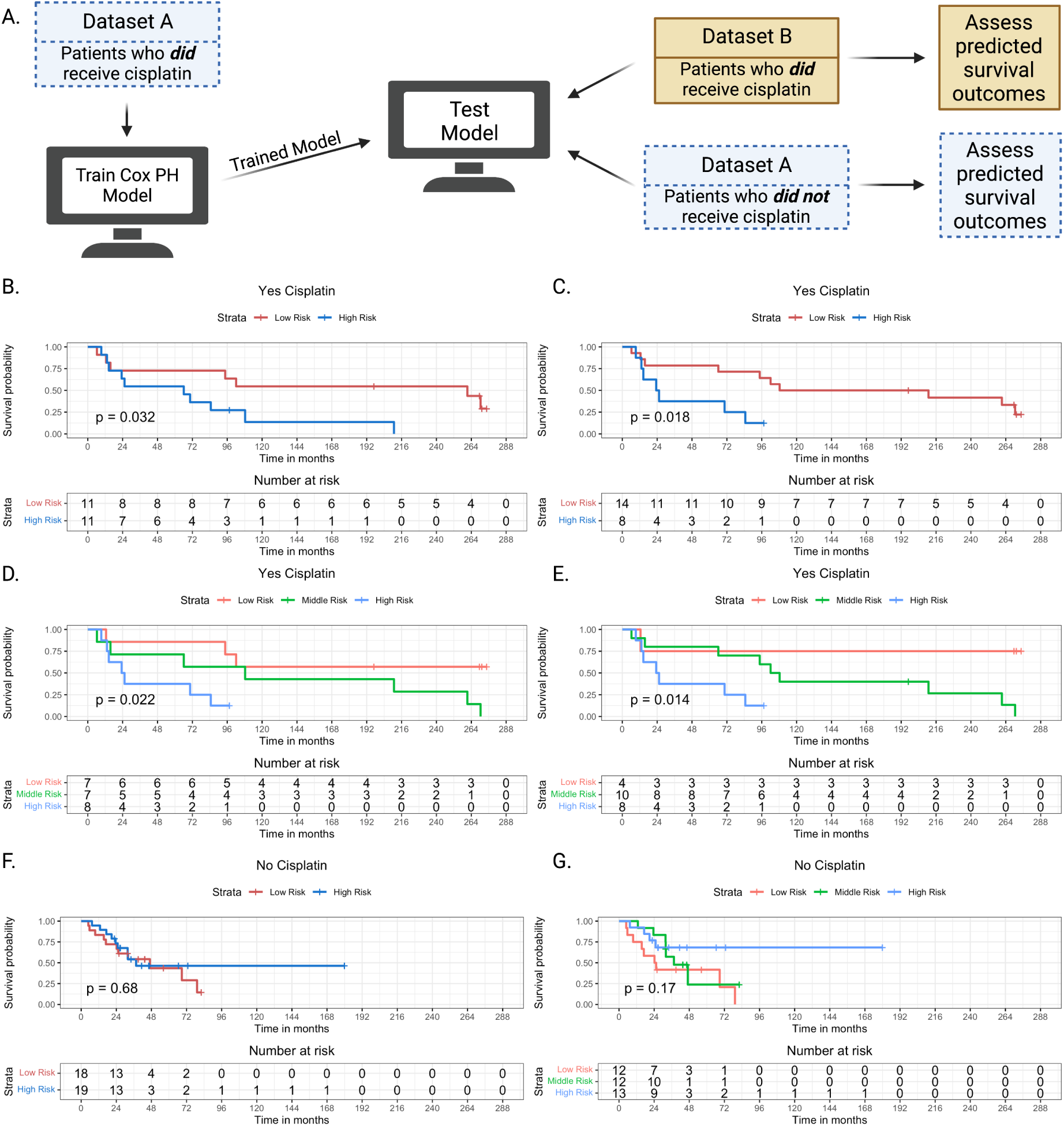
CisSig-trained model is predictive in patients who have received cisplatin, but lacks signal in patients who have not received cisplatin. A. Schematic description of model training and testing, where model is trained using patients who did receive cisplatin-containing treatment from Dataset A. Testing of the trained model is done using patients from the Dataset A who did not receive cisplatin-containing treatment and patients from the Dataset B who did receive cisplatin-containing treatment. B. Test samples that did receive cisplatin-containing treatment are separated into groups of “high” and “low risk” based on the model’s predictions using a median cutoff. Kaplan-meier curves show a significant separation between the two groups. C. The same analysis shown in B, using an optimal cutpoint (determined by chi-square statistic) instead of median to separate the cohorts. D-E. The same analyses shown in B-C, separating the groups into “high”, “middle”, and “low risk” groups using tertiles and the optimal two cutpoints, respectively. F-G. The same analyses shown in B and D, using samples from Dataset A that did not receive cisplatin-containing treatment, demonstrating no significant separation between the two groups.

## Discussion

The principles of convergent evolution tell us that genetically distant organisms can evolve similar traits in order to become more fit under the same selection pressure. In cancer, therefore, we cannot ignore the possibility that different mutations may lead to the same drug response phenotype. Therefore, our novel method groups convergent phenotypes and uses expression profiling to better predict drug response in cancer. In doing so, we harnessed the power of over 400 epithelial-origin cell lines in the GDSC Database to extract CisSig, a gene expression signature for use in predicting cisplatin response in epithelial-origin tumors.

Expression of CisSig provides knowledge that is different from and in addition to the presence of mutations in DNA damage response genes, Supplementary Figure 4. As demonstrated by many predictive modeling methods, our gene signature is highly effective at predicting drug response in GDSC cell lines. Yet, unlike with cell lines, high throughput characterization of drug response (i.e. IC50, AUC, etc) in clinical tumor samples is not feasible.^29^ Because of this, many researchers use survival as a surrogate measure of treatment response for tumor samples. However, without a known clinical history of cisplatin treatment, we cannot use survival as a surrogate measure of cisplatin response. Even in disease sites where there is level 1 evidence for use of cisplatin-containing chemotherapy (e.g. MIBC, triple-negative breast cancer), it cannot be assumed that all patients received this treatment, because many clinical factors may have prevented its use. Therefore, we assess CisSig’s translational capabilities across clinical datasets by demonstrating that increased expression of this signature is correlated to regular use of cisplatin among disease sites. In this analysis, GDSC was directly used in the extraction of CisSig and TCGA is used only for co-expression analysis in trimming the signature genes, but the TCC database was not used in any part of the extraction methodology.

Finally, we demonstrate that a CisSig-trained MIBC model can predict survival outcomes in a novel MIBC dataset for patients that received cisplatin-containing chemotherapy. Level 1 evidence for CisSig’s predictive capabilities in MIBC would would require validation in at least one additional cohort, but the results shown in Figure 6 show promising translational potential. Although there is a plethora of published gene expression data found on Gene Expression Omnibus, the lack of clinical annotation or use of targeted arrays makes additional clinical testing infeasible to the best knowledge of the authors. For example, many datasets contain pre-treatment samples from patients who later underwent cisplatin-containing chemotherapy and have publications that analyze survival outcomes for each patient, but the publicly available data do not include these outcomes (e.g. bladder: GSE87304; non-small cell lung cancer (NSCLC): GSE108492). Alternatively, there are some datasets that contain pre-treatment samples from patients who underwent cisplatin-containing chemotherapy, but the array used for gene expression profiling does not include all CisSig genes (e.g. ovarian: GSE23554; bladder: GSE5287; NSCLC: GSE14814).

Due to the empirical nature of our gene extraction method, the exact genes included in the final signature are of lower consequence than their combined predictive power. As such, we have not focused the validation of CisSig on the analysis of individual genes. It is, however, of note that the majority of the genes included in the signature are associated with tumorigenesis and tumor aggressiveness. Because cisplatin’s mechanism of action relies on disrupting actively replicating cells, it is not altogether surprising that increased expression of genes leading to cisplatin sensitivity would also promote poor prognosis in a treatment naïve setting. Furthermore, many of the genes have been denoted as possible therapeutic targets in a variety of epithelial-based cancers, such as *CDC7* in oral squamous carcinoma^30^ and liver cancer^31^, *ATP1B3* in gastric cancer^32^, and *FKBP14* in ovarian cancer.^33^ In a 2019 manuscript by Mucaki et al., the authors produce a cisplatin response signature from breast cancer cell lines in the GDSC dataset; however, their approach only includes genes with a known relationship to cisplatin response (none of which overlap with the genes in CisSig).^16^ Therefore, their final cisplatin response signature does not contain any CisSig genes. Another cisplatin response signature, extracted from 26 head and neck cancer patients with complete clinical response or non-response to cisplatin and 5-FU contains 10 genes that do not overlap with CisSig.^34^ Although these signatures do not show overlap with CisSig, they were both extracted from a specific disease site, while CisSig may be translated to a variety of disease sites. Finally, *CDC7A* in CisSig overlaps with the Mammaprint gene signature,^6^ and there are no overlapping genes with the OncotypeDx Breast Recurrence gene signature.^5^

This signature extraction method is, of course, not without limitations. First, a single tumor sample may not capture the intratumoral heterogeneity that is crucial for predicting the physiological response to a drug. Next, although the signature was extracted to find genes with importance across pan-cancer (epithelial-based) tumor subtypes, clinical validation must occur within individual disease sites. Given the heterogeneity between tumor subtypes, disease-site specific versions of CisSig may require trimming the genes of this pan-cancer signature even further, as seen with our preliminary analysis of CisSig in MIBC. Additionally, as discussed previously, using cell line expression data as the basis of a clinical signature is necessary given the current limitations of high throughput databases, but it can hinder translation. Therefore, a key future direction will be testing the signature in additional clinical data to determine if patient response to cisplatin can be stratified by signature expression.

Selection, such as drug treatment, acts on phenotype. And in this work, we demonstrate a novel gene signature extraction method–informed by principles of convergent evolution–where we find shared transcriptomic markers of drug response phenotype in tumors that appear genotypically disparate. By harnessing the power of a large dataset, such as the GDSC, we extracted a biologically-inspired product, CisSig. Expanding this method to produce signatures for response prediction to a variety of chemotherapeutic agents could lead to a monumental expansion of precision medicine in cancer.

## Methods

### Data Collection and Pre-Processing

All data cleaning, analysis, and plotting was performed using R (Version 4.0.5) with RStudio.

#### GDSC Gene Expression, Mutation, and Meta Data

Microarray mRNA expression, DNA mutation, drug response, and meta-data for 983 cell lines and 251 drugs was downloaded from the Genomics in Drug Sensitivity Database (GDSC)^35^. The expression, mutation, and meta-data were last updated 4 July 2016. The GDSC database can be accessed at https://www.cancerrxgene.org/. Documentation for the GDSC database states that the RMA normalized^36, 37^ expression data for all cell lines were collected via Human Genome U219 96-Array Plate using the Gene Titan MC instrument (Affymetrix). Further the robust multi-array analysis (RMA) algorithm was used to normalize the data, reporting intensity values for 18562 individual loci. The raw data and probe ID mappings were deposited in ArrayExpress (accession number: E-MTAB-3610). Whole exome sequencing was performed using the Agilent SureSelectXT Human All Exon 50Mb bait set. In our analysis, all protein coding mutations were labeled as having a “mutation present.” The RMA processed expression data and sequence variant (mutation) data is available at http://www.cancerrxgene.org/gdsc1000/.

Epithelial-based cell lines are extracted based on the following GDSC tissue descriptors (exact labels found in database): head and neck, oesophagus, breast, biliary_tract, large_intestine, liver, adrenal_gland, stomach, kidney, lung_NSCLC_adenocarcinoma, lung_NSCLC_squamous-_cell_carcinoma, mesothelioma, pancreas, skin_other, thyroid, Bladder, cervix, endometrium, ovary, prostate, testis, urogenital_system_other, uterus.

#### GDSC Drug Response Data

The drug response data in the GDSC database was last updated 27 March 2018; this version is referred to as “GDSC2.” Cisplatin drug concentration is reported in *µM*. Raw viability data were processed using the R package, gdscIC50, where they were normalized with negative controls (media alone) and positive controls (media only wells with no cells). Dose-response curves were fit using a multi-level fixed effect model with a classic sigmoidal curve shape assumed. This model was fitted using all cell line/drug combinations that were screened instead of fitting separate models to individual drug-response series. In this approach, the shape parameter only changes between cell lines, but the position parameter is adjusted between cell lines and compounds. Additional information regarding dose-response curve fitting may be found at Vis et al.^38^. Fitting models to all dose-response series leads to improved robustness for more accurate IC50 and AUC estimates.

#### TCGA Gene Expression Data

RNA-Seq by Expectation Maximization (RSEM) normalized gene expression for epithelial-based cancers was downloaded from The Cancer Genome Atlas (TCGA) database, which was accessed through the Firebrowse database using the ‘RTCGAToolbox’ package (version 2.20.0)^39^ in R. The following TCGA Study Abbreviations were downloaded (exact labels found in database): ACC, BLCA, BRCA, CESC, CHOL, COADREAD, ESCA, HNSC, KIRC, KIRP, KICH, LIHC, LUAD, LUSC, MESO, OV, PAAD, PRAD, STAD, THCA, THYM, UCEC. These values were measured through the Illumina HiSeq RNAseq V2 platform and were log2 transformed.

#### Total Cancer Care (TCC) Gene Expression Data

The Total Cancer Care Dataset is collected by the H. Lee Moffitt Cancer Center and Research Institute using protocols described in Fenstermacher et al^40, 41^. The Total Cancer Care (TCC) protocol is a prospective tissue collection protocol that has been active at Moffitt Cancer Center (Tampa, FL, USA) and 17 other institutions since 2006. We assayed tumours from adult patients enrolled in the TCC protocol on Affymetrix Hu-RSTA-2a520709, which contains approximately 60,000 probesets representing 25,000 genes. Chips were normalised using iterative rank-order normalisation.^42^ Batch effects were reduced using partial-least squares. We extracted from the TCC database normalised, debatched expression values for 9,063 samples from 17 sites of epithelial origin and the 19 CisSig genes. We excluded all metastatic duplicate samples and disease sites with fewer than 25 samples.

### Drug Response Quality Control

IC50 is an imperfect measure of drug response, yet it is widely used throughout the literature. It is defined as the concentration of drug at which cells experience 50% inhibitory effect. Another measure of drug response is area under the drug response curve, which is defined as the integral of a drug response curve, where cellular activity is measured on the y-axis and drug concentration is measured on the x-axis. IC50 and AUC values for all epithelial cell lines are compared using a Spearman correlation test (see **Supplementary Figure 1**) in order to assess concordance between the two metrics.

### Differential Gene Expression Analysis

As seen in **Figure 2C**, the GDSC dataset is split into 5-folds, where 20% of the cell lines are removed from further analysis for each of the 5 runs. This leaves 343 or 344 cell lines in each of the 5 partitions. After data partitioning, the top 20% and bottom 20% are extracted for comparison using differential expression analysis, **Figure 2C**.

Differential expression analysis is performed using three algorithms: significance analysis of microarrays (SAM), resampling-based multiple hypothesis testing, and linear models for microarrays (limma), which are implemented using R packages ‘samr’^24^ (version 3.0), ‘multtest’^25^ (version 2.46.0), and ‘limma’^23^ (version version 3.46.0), respectively. Gene expression was pre-normalized using RMA (discussed above) and genes were not pre-filtered before this analysis. This analysis has 69 samples per group, which is appropriate given the demonstration by Baccarella et al. showing that differential expression results begin to vary problematically beginning when there are as few as 8 samples per group^43^.

A false discovery rate or p-value cutoff of 0.20 was chosen for each method. The ‘samr’ and ‘multtest’ method were both set to the same seed. The ‘samr’ method used 10,000 permutations (parameter: “nperm”) and test statistic was set to “standard” for t-test (parameter: “testStatistic”). The ‘limma’ method used no p-value adjustment method (parameter: “adjust.method”) and a log-fold change cutoff of 0.5 (parameter: “lfc”). The ‘multtest’ method used 1,000 bootstrap iterations (parameter: “B”) and single-step minP for multiple testing procedure (parameter: “method”). All other parameters for the three algorithms were set to default. The intersection of the genes found to have significantly increased expression in sensitive cell lines by the three algorithms is termed “seed genes” for use in future co-expression analysis. An FDR cutoff of 0.2 is a relatively non-stringent FDR cutoff; it was chosen in order to include a variety of genes before taking the intersection of results between the three methods.

### Co-Expression Network Analysis and Final Signature Derivation

The co-expression network, represented in the pipeline of **Figure 2B**, is made by performing a pairwise Spearman correlation between the expression of each seed gene and every other gene (including other seed genes) except itself. The correlation coefficient for each pairwise comparison is termed the “affinity score.” Next, the network is transformed so that the largest 5% of affinity scores are transformed to 1 and all other scores become 0. This is done without squaring the scores in order to extract only positive correlations. The average affinity score for each gene compared to each seed gene is then derived; this value becomes known as a gene’s “connectivity score.” The intersection between the differentially expressed seed genes and genes with the top 20% of the highest connectivity scores become known as the “connectivity genes.” Five sets of connectivity genes are compiled, one for each data partition. The final signature (CisSig) is produced by extracting any gene that is found in at least three of the five connectivity gene sets.

### Signature Quality Control in TCGA

In order to examine how CisSig compares to the original differential gene expression results and ensure portability to novel datasets, we perform a quality control analysis within the TCGA dataset using the ‘sigQC’ R package^26^ with methodology as in Dhawan et al. 2019 ^27^. Here, various metrics are calculated using the expression of the genes found in the gene expression signature and the 5 sets of differential expression analysis results. These metrics include intra-signature correlation, correlation between the mean expression and first principal component, and skewness of the signature expression. The final results of all the metrics calculated for each signature are displayed in a radar plot, with a summary score of each set of genes (signature) tested. This summary score is the ratio of the area within the radar plot and the full polygon if each metric was the highest value possible.

### Predicting cell line IC50 using CisSig in GDSC

A cell line or sample’s median normalized expression value of the CisSig genes is termed the CisSig score. Cell lines were again organized into five folds (independent of the data partitioning used in the signature extraction, described in **Figure 2C**). Predictive models were built using 80% of the cell lines (training cell lines) and tested on the 20% of the cell lines withheld from the model (validation cell lines). All models were built with two versions of input–one using all of the epithelial-based cell lines in the GDSC database and the other using only the cell lines in the top and bottom quintiles of CisSig score. When using all the epithelial-based cell lines, training sets consist of 344-345 cell lines, while testing sets consist of 86 cell lines. When using only the cell lines in the top and bottom quintiles for signature expression, training sets consist of 137 or 138 cell lines and testing sets consist of 34 or 35 cell lines.

Simple linear and logistic regression was used to predict IC50 as a continuous variable with CisSig score as the input. Elastic net, L1-, and L2-penalized linear regression methods utilized the expression of each of the 19 CisSig genes to predict IC50 as a continuous variable. Elastic net, L1-, and L2-penalized logistic regression methods, support vector machine (SVM), and random forest methods utilized expression of each of the 19 CisSig genes to predict IC50 as a binary variable (above or below the median of the group). All linear regression models were evaluated using the Spearman correlation coefficient between true and predicted IC50 values from the validation set. Classification models (logistic regression, SVM, and random forest) were evaluated using area under the receiver operating characteristic (ROC) curve (AUC).

Elastic net, L1-, and L2-penalized linear and logistic regression models were built using the ‘glmnet’ package (version 4.1-2) in R. The alpha parameter was set to 0.5, 1, and 0 for elastic net, L1-, and L2-penalized regression, respectively. Models were tuned with 10-fold cross validation to choose a value for lambda with the best predictive capabilities based on mean square error for linear models and misclassification error for logistic models.

SVM models were built with the ‘e1071‘ package (version 1.7-8) in R, using both a linear and polynomial kernel. Models were tuned with 10-fold cross validation to choose the best value for degree (from 3, 4, 5), gamma (from 10^-3^, 10^-2^, 10^-1^, 1, 10^1^, 10^2^, 10^3^), and cost (from 10^-3^, 10^-2^, 10^-1^, 1, 10^1^, 10^2^, 10^3^).

Random forest models were built with the ‘randomForest’ package (version 4.6-14), and each model grew 500 trees. All other parameters in training the prediction models were default.

### Cell Line Persistence Curves

Cell lines with high CisSig scores (predicting the more sensitive cell lines) and low signatures scores (predicting the more resistant cell lines) are separated by quintile. A Kaplan-Meier survival model is built for the two cohorts using IC50 in lieu of survival time, using the ‘Surv‘ and ‘survfit‘ function from the ‘survival‘ R package and ‘ggsurvplot‘ from the ‘survminer‘ R package. A log-rank test (‘ggsurvplot‘ from the ‘survminer‘ R package) compares the two survival curves to analyze if the two cohorts of signature expression are related to different “survival” of higher IC50s in each group.

### Null distributions of cell line IC50 models

CisSig’s performance was compared to a null distribution for all models built, including all models used to predict IC50 as a continuous or binary variable and the cell line persistence models using the log-rank test to compare the two survival curves. To build each null distribution, 1000 random gene signatures with the same length as CisSig were chosen. Each random gene signature was selected using all genes included in the GDSC expression profiling without replacement. The performance of each random signature was tested in each individual modeling method, producing a null distribution for each modeling method. As discussed above, the predictive models utilize five-fold cross validation and the best summary statistic of the five folds is chosen to represent the signature’s performance. This remains consistent for the null models, where the best summary statistic of the five folds is used to represent each random signature. Again, all code for building the testing and null models may be found in the GitHub repository listed in Code and Data Availability.

### Comparing mutation status and CisSig Score

Epithelial cell lines were separated into high and low cohorts based on being above or below the median CisSig Score. For each of the 17 DNA damage response genes shown in Supplementary Figure 4, a chi-square test compared the presence of a mutation in cell lines with high or low CisSig score. This was performed using the ‘chisq.test‘ function in the ‘stats‘ package in R. P-values underwent Bonferroni correction.

### Ranking disease sites in GDSC, TCGA, and TCC by CisSig Score

All epithelial-origin cell lines or tumor samples in the GDSC, TCGA, and TCC datasets had CisSig Score calculated as previously described. For the purposes of plotting on a log-scale, the scores were linearly adjusted by adding the absolute value of the lowest score plus 1 to each sample’s score, making the lowest score now 1. For example, if the lowest signature score for the dataset was -5, 6 was added to each sample’s score. Disease sites within each dataset were ranked by median CisSig score. For disease sites shared between datasets, a Spearman correlation was performed to assess how the rank of disease sites compare between datasets.

### Classifying disease sites by cisplatin use

NCCN Treatment Guidelines for each disease site were manually searched, versions listed in **Supplementary Table 8**. Disease sites were classified as including cisplatin in treatment guidelines, only including cisplatin in very select circumstances, or not including cisplatin in treatment guidelines. For those classified as only using cisplatin in select circumstances, details are noted in **Supplementary Table 8**.

### Survival analysis in external MIBC cohorts

Two separate models were trained, using a similar method displayed in Figure 6A and Supplementary Figure 16A, respectively. For the model trained in Figure 6A, we performed univariate analysis for each CisSig gene to predict overall survival of samples that received cisplatin-containing chemotherapy in Dataset A. CisSig genes with a variance value of *<* 0.2 using the ‘var‘ function in the ‘stats‘ package were not included in this analysis. A multivariate model was trained using genes from the univariate analysis that demonstrated a coefficient of *−*0.5 or lower; this became the trained model. Both univariate and multivariate models were build using the ‘Surv‘ and ‘coxph‘ function from the ‘survival‘ package in R. The trained model was tested using the ‘predict‘ function in R, extracting the linear predictor for samples from Dataset B who received cisplatin-containing neoadjuvant chemotherapy and samples from Dataset A who did not receive any cisplatin-containing treatment. Samples were separated by median, optimal single cutpoint, tertiles, and optimal double cutpoints. Cohorts separated by each cutpoint were compared using Kaplan-Meier analysis, using the ‘ggsurvplot‘ function in the ‘survminer‘ package in R. The same analysis was performed for Supplementary Figure 16, except the training dataset was Dataset B (patients who received cisplatin-containing neoadjuvant chemotherapy), while the testing datasets were patients from Dataset A who did and did not receive cisplatin-containing neoadjuvant chemotherapy. The optimal cutpoints for separating cohorts in the Kaplan-Meier analyses found in Figure 6C and E and Supplementary Figure 16C and E are found by searching all possible cutpoints where each cohort has at least 4 patients, selecting for which cutpoint leads to the greatest chi-square statistic.

### Data and code availability

The code to download all data, extract CisSig, perform validation of the signature, and reproduce all figures in the manuscript is available via GitHub at https://github.com/jessicascarborough/cissig. .

### Statistics Study Approval

The Genomics of Drug Sensitivity in Cancer (GDSC) dataset was accessed using R (code in the cited GitHub repository) to directly download files from the following links, which do not require registration: https://www.cancerrxgene.org/gdsc1000/GDSC1000_WebResources//Data/preprocessed/Cell_line_RMA_proc_basalExp.txt.zip, ftp://ftp.sanger.ac.uk/pub/project/cancerrxgene/releases/current_release/GDSC2_fitted_dose_response_25Feb20.xlsx, ftp://ftp.sanger.ac.uk/pub/project/cancerrxgene/releases/current_release/Cell_Lines_Details.xlsx. The Cancer Genome Atlas (TCGA) dataset was accessed using the ‘RTCGAToolbox‘ R package (code in the cided GitHub repository) to download each disease site’s RSEM normalized RNASeq V2 (labeled ‘RNASeq2GeneNorm‘). The data can be accessed, without registration here: https://gdac.broadinstitute.org/. The Total Cancer Care (TCC) dataset requires application for access, and can be found here: https://moffitt.org/research-science/total-cancer-care/. Approval was received to use the anonymized data in this manuscript after registration, and the IRB of Moffitt Cancer Center gave ethical approval for the collection of the original data.

## Data Availability

All data produced can be accessed by following code available online at https://github.com/jessicascarborough/cissig

## Author contributions

J.A.S. contributed to experimental design, wrote all associated code, analyzed data, and wrote the manuscript. S.A.E. analyzed data. J.T.R. contributed to experimental design. J.G.S. contributed to experimental design, analyzed data, and wrote the manuscript. A.D. contributed to experimental design, analyzed data, and wrote the manuscript. All authors read and approved of the manuscript.

## Acknowledgements

would like to thank NIH (5R37CA244613-02) and the American Cancer Society (RSG-20-096-01) for their generous support. J.A.S. thanks the NIH for their support through the T32GM007250 and 1F30CA257076-01 grants.

The results published here are in whole or part based upon data generated by the TCGA Research Network: https://www.cancer.gov/tcga. All authors are grateful to the cancer patients who provided tissue for further study in the GDSC, TCGA, and TCC datasets.

This work made use of the High Performance Computing Resource in the Core Facility for Advanced Research Computing at Case Western Reserve University.

Figures 1-6 and Supplementary Figures 2, 4, and 16 were made in part or in whole using BioRender.com.

## Supplementary Tables

**Table S1.**
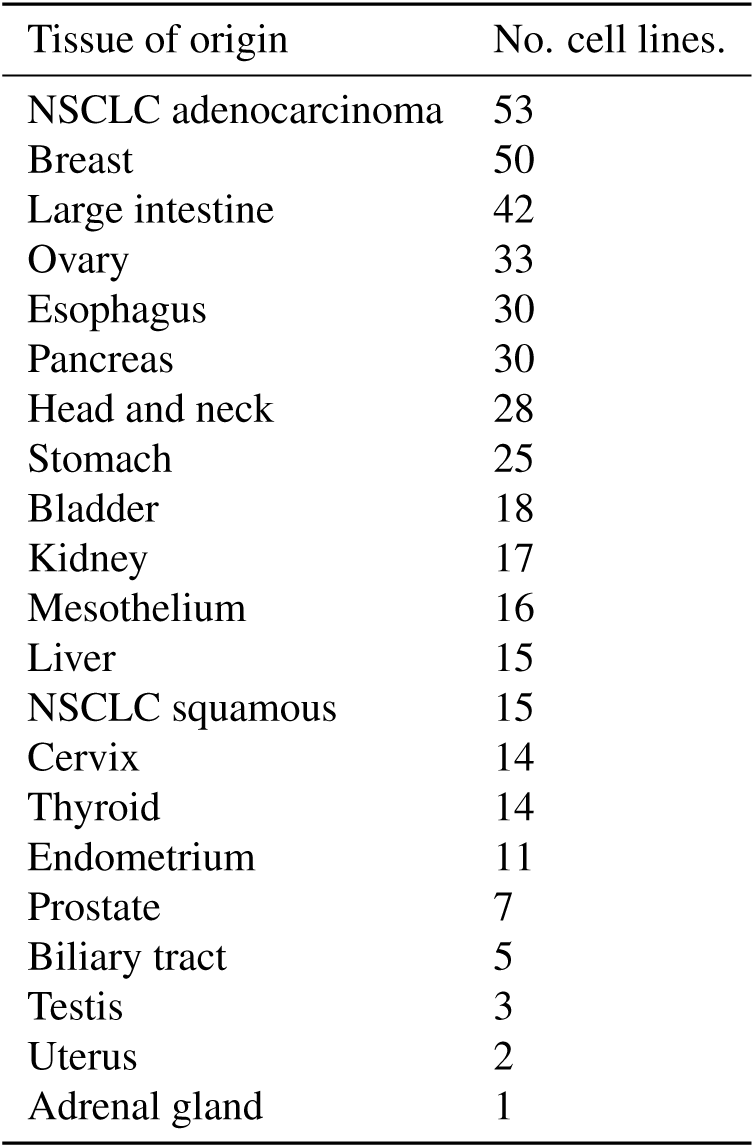
Tissue of origin for all 429 epithelial-origin GDSC cell lines.

| Tissue of origin | No. cell lines. |
| --- | --- |
| NSCLC adenocarcinoma | 53 |
| Breast | 50 |
| Large intestine | 42 |
| Ovary | 33 |
| Esophagus | 30 |
| Pancreas | 30 |
| Head and neck | 28 |
| Stomach | 25 |
| Bladder | 18 |
| Kidney | 17 |
| Mesothelium | 16 |
| Liver | 15 |
| NSCLC squamous | 15 |
| Cervix | 14 |
| Thyroid | 14 |
| Endometrium | 11 |
| Prostate | 7 |
| Biliary tract | 5 |
| Testis | 3 |
| Uterus | 2 |
| Adrenal gland | 1 |

**Table S2.**
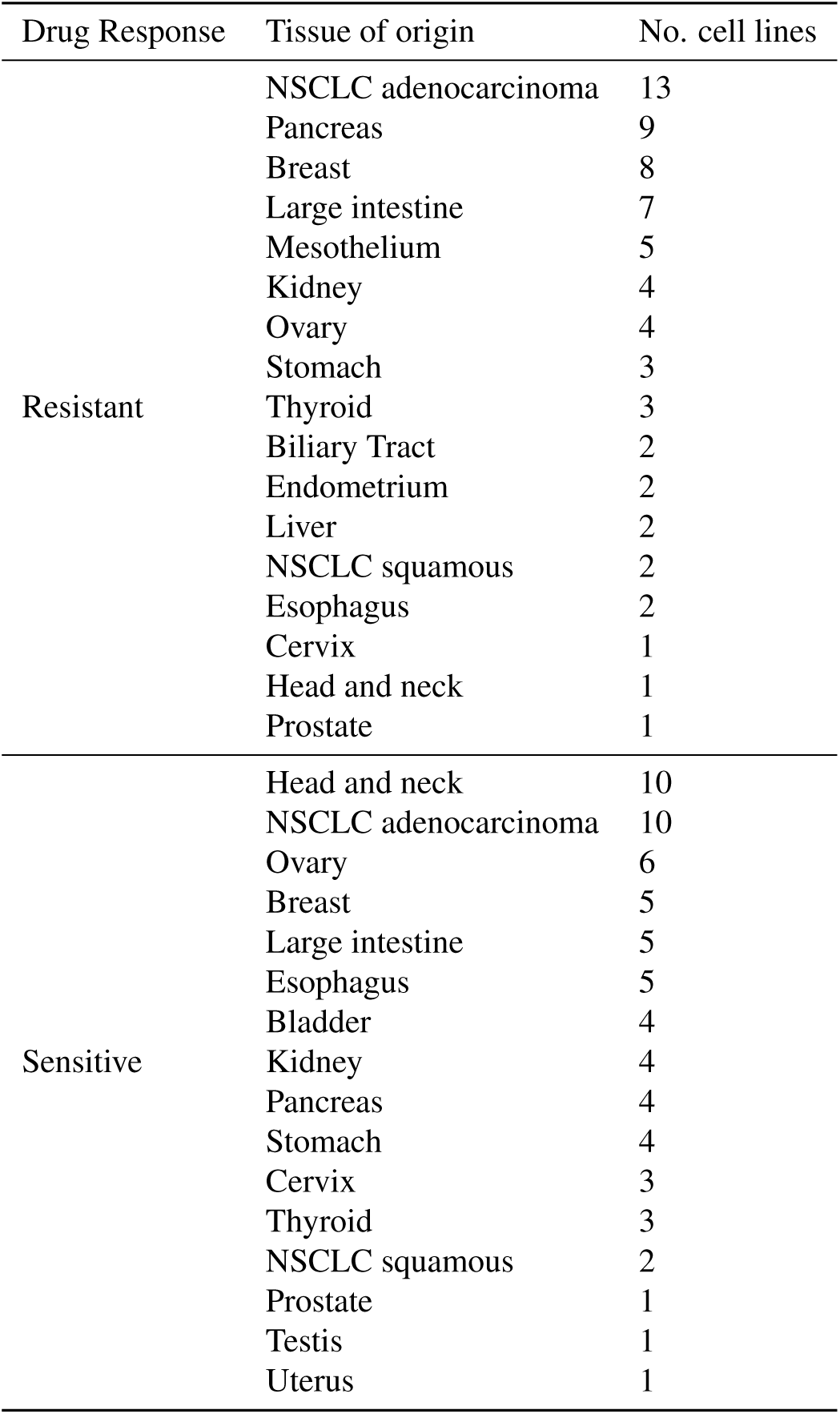
Tissue of origin for DE comparison groups for fold 1.

| Drug Response | Tissue of origin | No. cell lines |
| --- | --- | --- |
| Resistant | NSCLC adenocarcinoma | 13 |
|  | Pancreas | 9 |
|  | Breast | 8 |
|  | Large intestine | 7 |
|  | Mesothelium | 5 |
|  | Kidney | 4 |
|  | Ovary | 4 |
|  | Stomach | 3 |
|  | Thyroid | 3 |
|  | Biliary Tract | 2 |
|  | Endometrium | 2 |
|  | Liver | 2 |
|  | NSCLC squamous | 2 |
|  | Esophagus | 2 |
|  | Cervix | 1 |
|  | Head and neck | 1 |
|  | Prostate | 1 |
| Sensitive | Head and neck | 10 |
|  | NSCLC adenocarcinoma | 10 |
|  | Ovary | 6 |
|  | Breast | 5 |
|  | Large intestine | 5 |
|  | Esophagus | 5 |
|  | Bladder | 4 |
|  | Kidney | 4 |
|  | Pancreas | 4 |
|  | Stomach | 4 |
|  | Cervix | 3 |
|  | Thyroid | 3 |
|  | NSCLC squamous | 2 |
|  | Prostate | 1 |
|  | Testis | 1 |
|  | Uterus | 1 |

**Table S3.** Tissue of origin for DE comparison groups for fold 2.

| Drug Response | Tissue of origin | No. cell lines |
| --- | --- | --- |
| Resistant | NSCLC adenocarcinoma | 13 |
|  | Breast | 9 |
|  | Pancreas | 9 |
|  | Large intestine | 5 |
|  | Mesothelium | 5 |
|  | Stomach | 5 |
|  | Esophagus | 4 |
|  | Ovary | 4 |
|  | Biliary Tract | 2 |
|  | Head and neck | 2 |
|  | Kidney | 2 |
|  | Liver | 2 |
|  | NSCLC squamous | 2 |
|  | Thyroid | 2 |
|  | Bladder | 1 |
|  | Endometrium | 1 |
|  | Prostate | 1 |
| Sensitive | Head and neck | 11 |
|  | NSCLC adenocarcinoma | 8 |
|  | Bladder | 6 |
|  | Breast | 5 |
|  | Large intestine | 5 |
|  | Esophagus | 5 |
|  | Ovary | 5 |
|  | Stomach | 5 |
|  | Cervix | 3 |
|  | Kidney | 3 |
|  | Pancreas | 3 |
|  | NSCLC squamous | 2 |
|  | Testis | 2 |
|  | Thyroid | 2 |
|  | Endometrium | 1 |
|  | Prostate | 1 |
|  | Uterus | 1 |

**Table S4.** Tissue of origin for DE comparison groups for fold 3.

| Drug Response | Tissue of origin | No. cell lines |
| --- | --- | --- |
| Resistant | NSCLC adenocarcinoma | 13 |
|  | Pancreas | 10 |
|  | Breast | 8 |
|  | Large intestine | 5 |
|  | Mesothelium | 5 |
|  | Kidney | 4 |
|  | Ovary | 4 |
|  | Stomach | 4 |
|  | Esophagus | 3 |
|  | Thyroid | 3 |
|  | Endometrium | 2 |
|  | Head and neck | 2 |
|  | Biliary Tract | 1 |
|  | Bladder | 1 |
|  | Cervix | 1 |
|  | Liver | 1 |
|  | NSCLC squamous | 1 |
|  | Prostate | 1 |
| Sensitive | Ovary | 10 |
|  | Head and neck | 9 |
|  | Bladder | 8 |
|  | NSCLC adenocarcinoma | 6 |
|  | Esophagus | 5 |
|  | Breast | 4 |
|  | Cervix | 4 |
|  | Kidney | 4 |
|  | Pancreas | 4 |
|  | Stomach | 4 |
|  | Large intestine | 3 |
|  | Thyroid | 3 |
|  | Testis | 2 |
|  | Endometrium | 1 |
|  | NSCLC squamous | 1 |
|  | Uterus | 1 |

**Table S5.** Tissue of origin for DE comparison groups for fold 4.

| Drug Response | Tissue of origin | No. cell lines |
| --- | --- | --- |
| Resistant | NSCLC adenocarcinoma | 12 |
|  | Breast | 10 |
|  | Large intestine | 8 |
|  | Pancreas | 6 |
|  | Mesothelium | 5 |
|  | Ovary | 5 |
|  | Thyroid | 4 |
|  | Kidney | 3 |
|  | Liver | 3 |
|  | Esophagus | 3 |
|  | Biliary Tract | 2 |
|  | Head and neck | 2 |
|  | Stomach | 2 |
|  | Bladder | 1 |
|  | Cervix | 1 |
|  | Endometrium | 1 |
|  | NSCLC squamous | 1 |
| Sensitive | Head and neck | 9 |
|  | NSCLC adenocarcinoma | 8 |
|  | Ovary | 8 |
|  | Bladder | 6 |
|  | Kidney | 5 |
|  | Large intestine | 5 |
|  | Stomach | 5 |
|  | Cervix | 4 |
|  | Esophagus | 4 |
|  | Pancreas | 3 |
|  | Thyroid | 3 |
|  | Breast | 2 |
|  | Testis | 2 |
|  | Endometrium | 1 |
|  | NSCLC squamous | 1 |
|  | Prostate | 1 |
|  | Uterus | 1 |

**Table S6.**
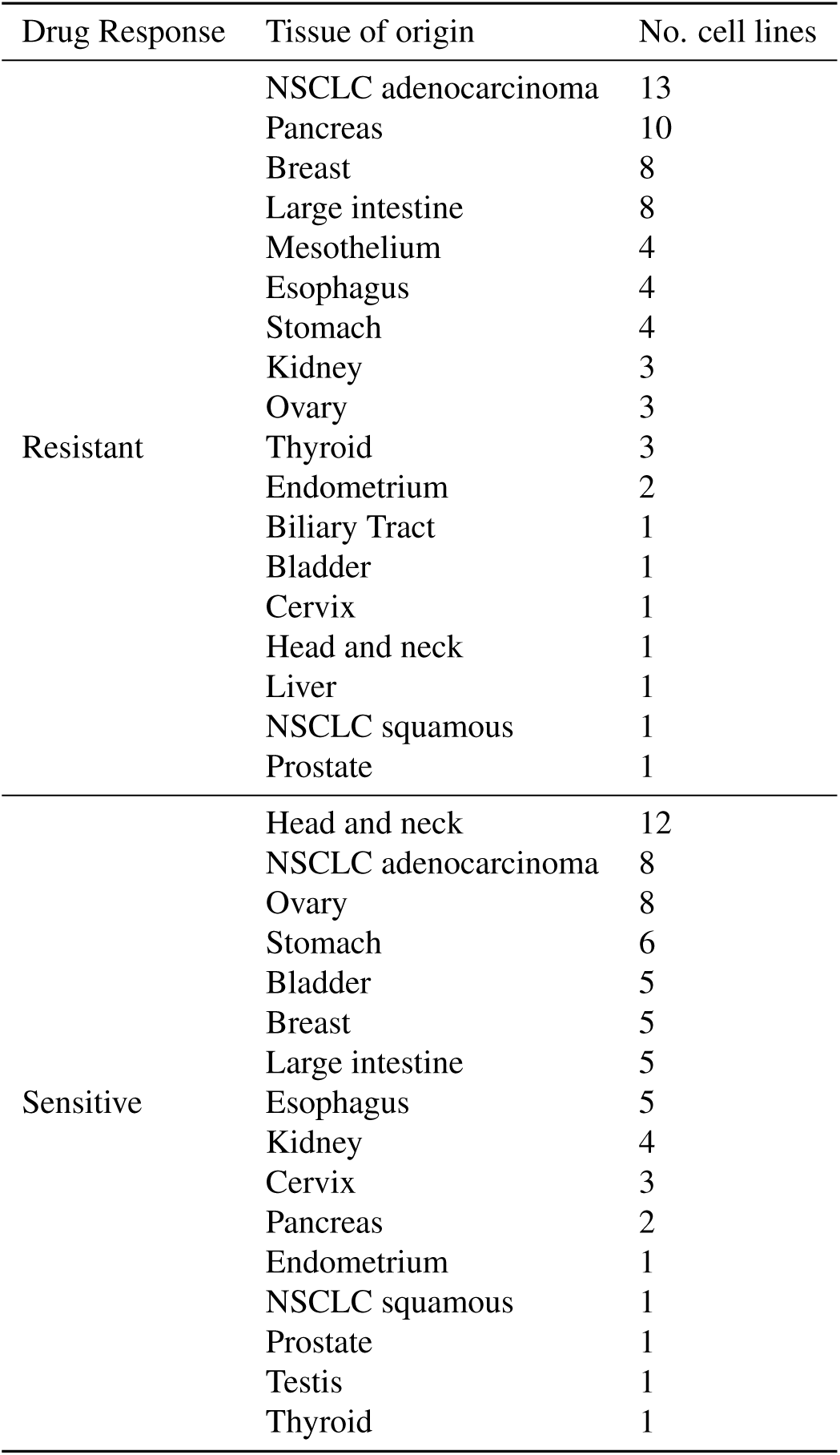
Tissue of origin for DE comparison groups for fold 5.

**Table S7.** DE genes by fold. The SAM method consistently extracts more genes than limma or multtest. The intersection, however, is much smaller than either limma or multtest, showing significant filtering during the intersection step.

| Fold | Method | No. Up-regulated Genes | No. Down-regulated Genes |
| --- | --- | --- | --- |
| 1 | SAM | 1979 | 1083 |
|  | limma | 181 | 322 |
|  | multtest | 219 | 150 |
|  | intersection | 59 | 58 |
| 2 | SAM | 1397 | 853 |
|  | limma | 159 | 302 |
|  | multtest | 139 | 115 |
|  | intersection | 32 | 41 |
| 3 | SAM | 2290 | 1143 |
|  | limma | 176 | 355 |
|  | multtest | 247 | 173 |
|  | intersection | 58 | 73 |
| 4 | SAM | 1904 | 1069 |
|  | limma | 188 | 263 |
|  | multtest | 237 | 147 |
|  | intersection | 61 | 42 |
| 5 | SAM | 566 | 636 |
|  | limma | 156 | 221 |
|  | multtest | 93 | 87 |
|  | intersection | 34 | 28 |

**Table S8.**
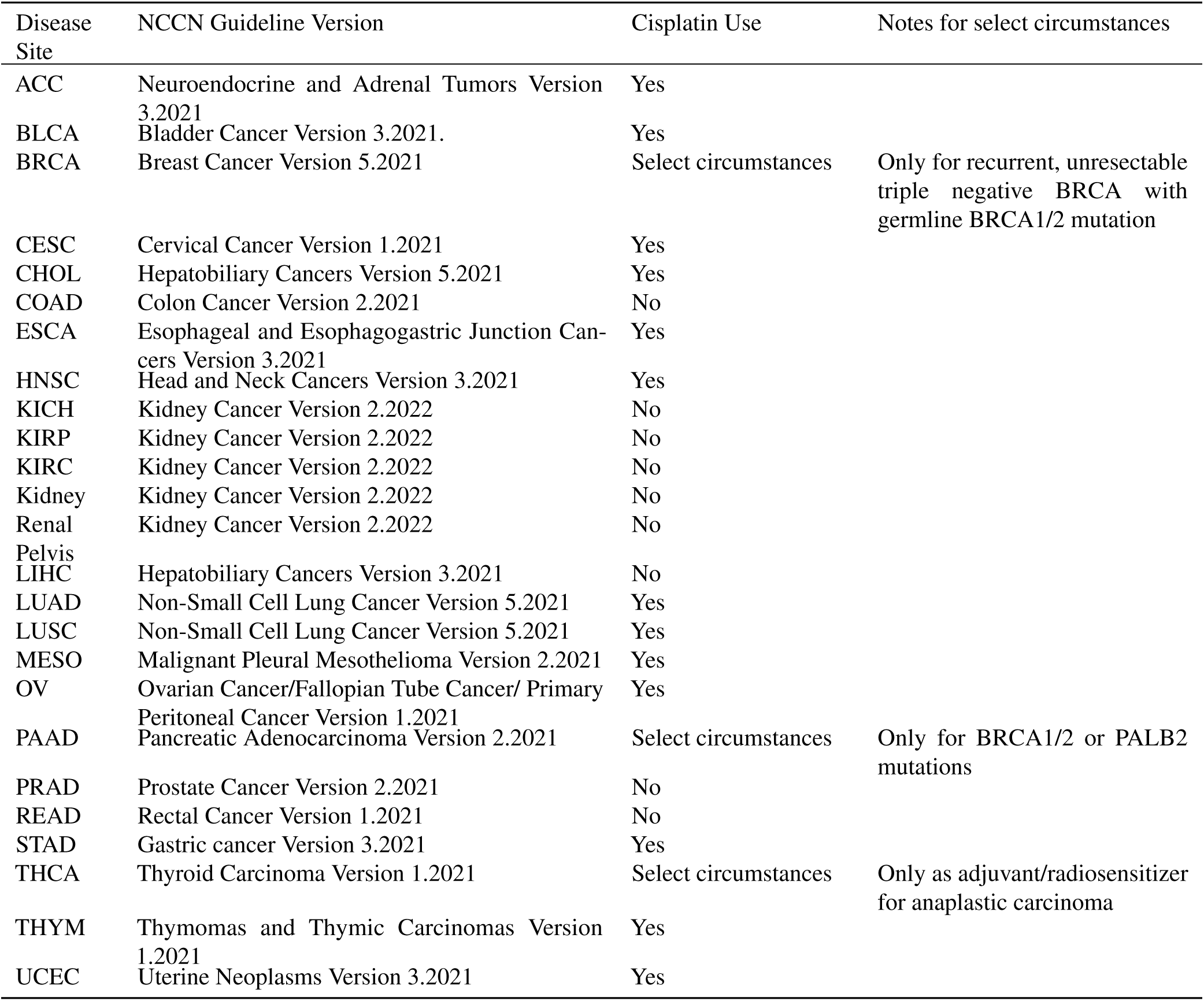
NCCN Guideline versions used for assessing disease-site specific treatment guidelines.

**Table S9.** Resulting coefficients for multivariate model trained in Figure 6.

| Gene | Coefficient Estimate | Exponentiated Coefficient Estimate | Coefficient Standard Error |
| --- | --- | --- | --- |
| C15orf41 | -2.2185 | 0.1088 | 1.3481 |
| FKBP14 | -1.5948 | 0.2029 | 1.0083 |
| PSAT1 | -1.8362 | 0.1594 | 1.5889 |
| C1QBP | 0.2797 | 1.3227 | 1.0234 |

**Table S10.**
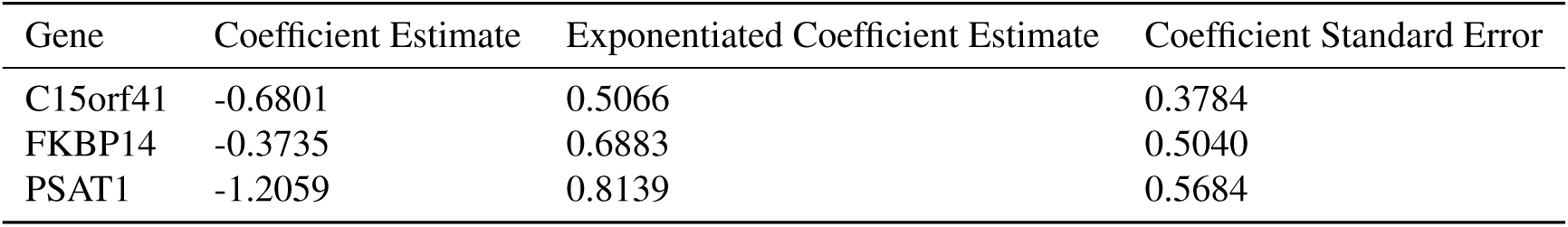
Resulting coefficients for multivariate model trained in **Supplementary Figure 16**.

## Supplementary Figures

**Figure S1.**
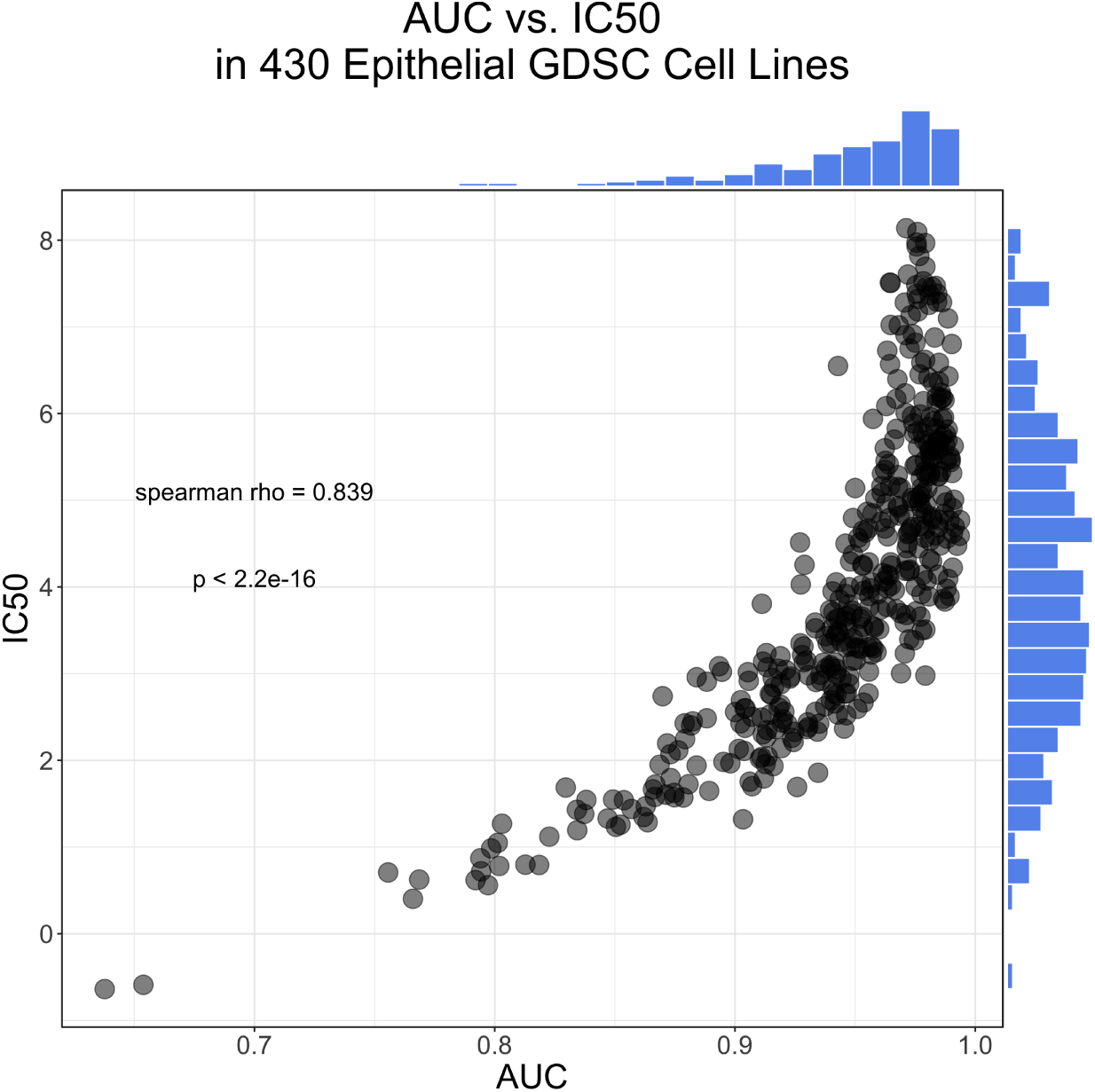
Correlation between AUC and IC50 drug response metrics for epithelial-based cancer cell lines in the Genomics of Drug Discovery in Cancer (GDSC) dataset.

**Figure S2.**
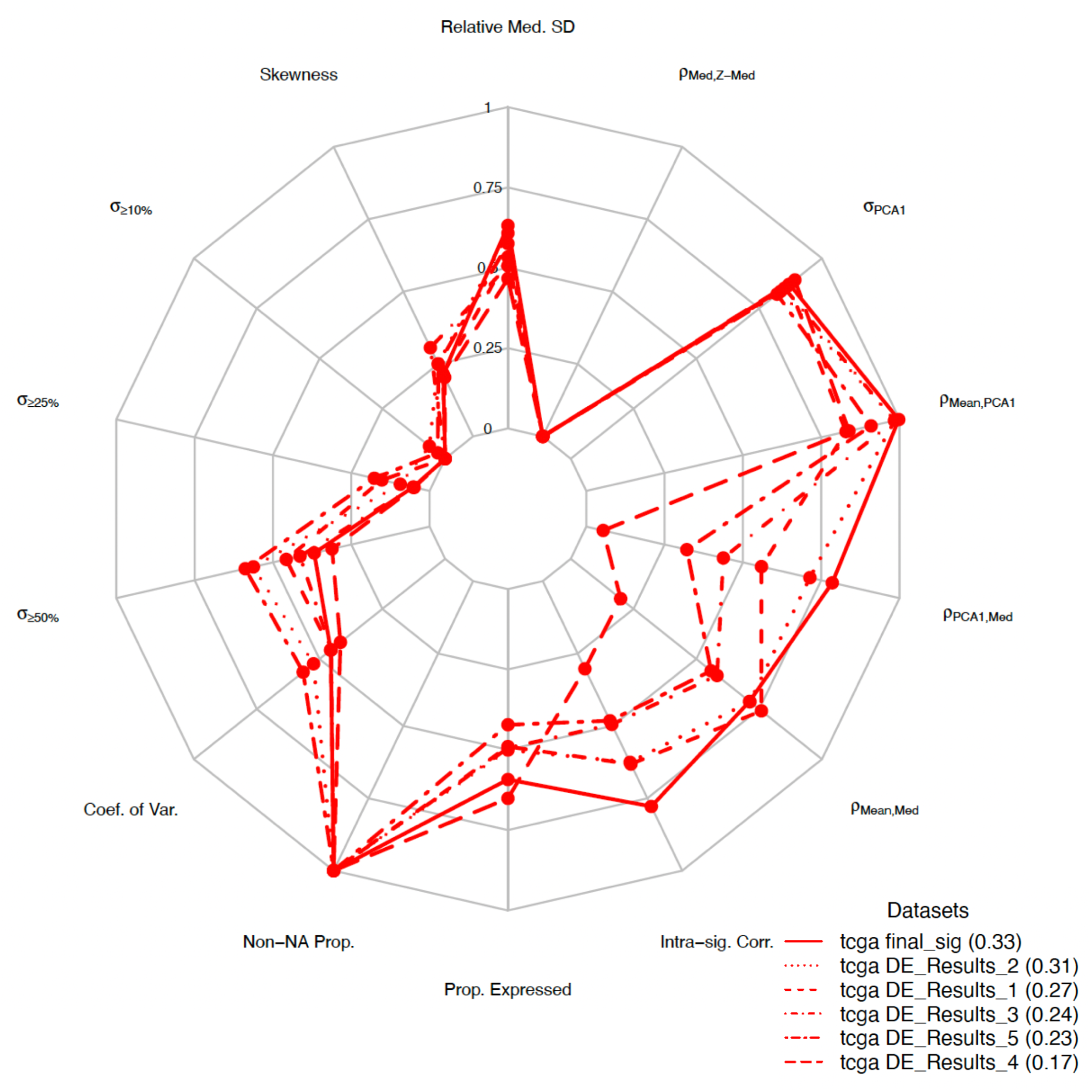
**Quality control metrics comparing differential expression results to the final gene signature using sigQC**^26, 27^. CisSig is compared to the folds of differential gene expression analysis, comparing results using a radar plot. It shows greater intra-signature correlation, higher correlation between signature mean and median, and decreased skewness within RNA-seq expression from TCGA samples of epithelial origin.

**Figure S3.**
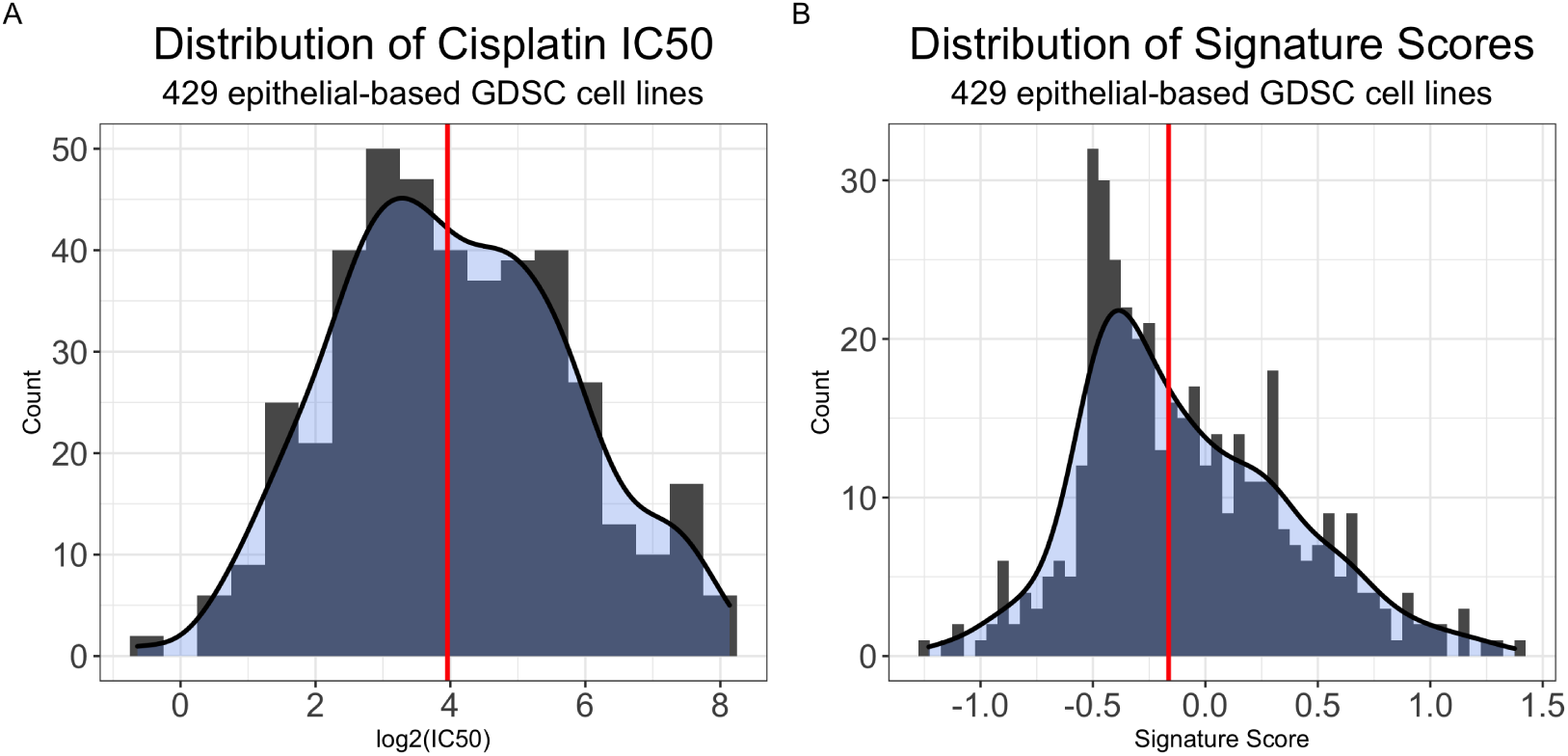
Cisplatin IC50 (log2-transformed) in epithelial-origin GDSC cell lines is relatively normally distributed, while CisSig Score has a slight right skew. A. Distribution of CisSig across 429 epithelial-based GDSC cell lines, using a histogram (gray) and kernel density estimation (blue). Median score marked by red vertical line. CisSig score is calculated as a cell line’s median normalized expression of CisSig genes listed in A. B. Distribution of cisplatin IC50 across 429 epithelial-based GDSC cell lines, using a histogram (gray) and kernel density estimation (blue). Median IC50 marked by red vertical line.

**Figure S4.**
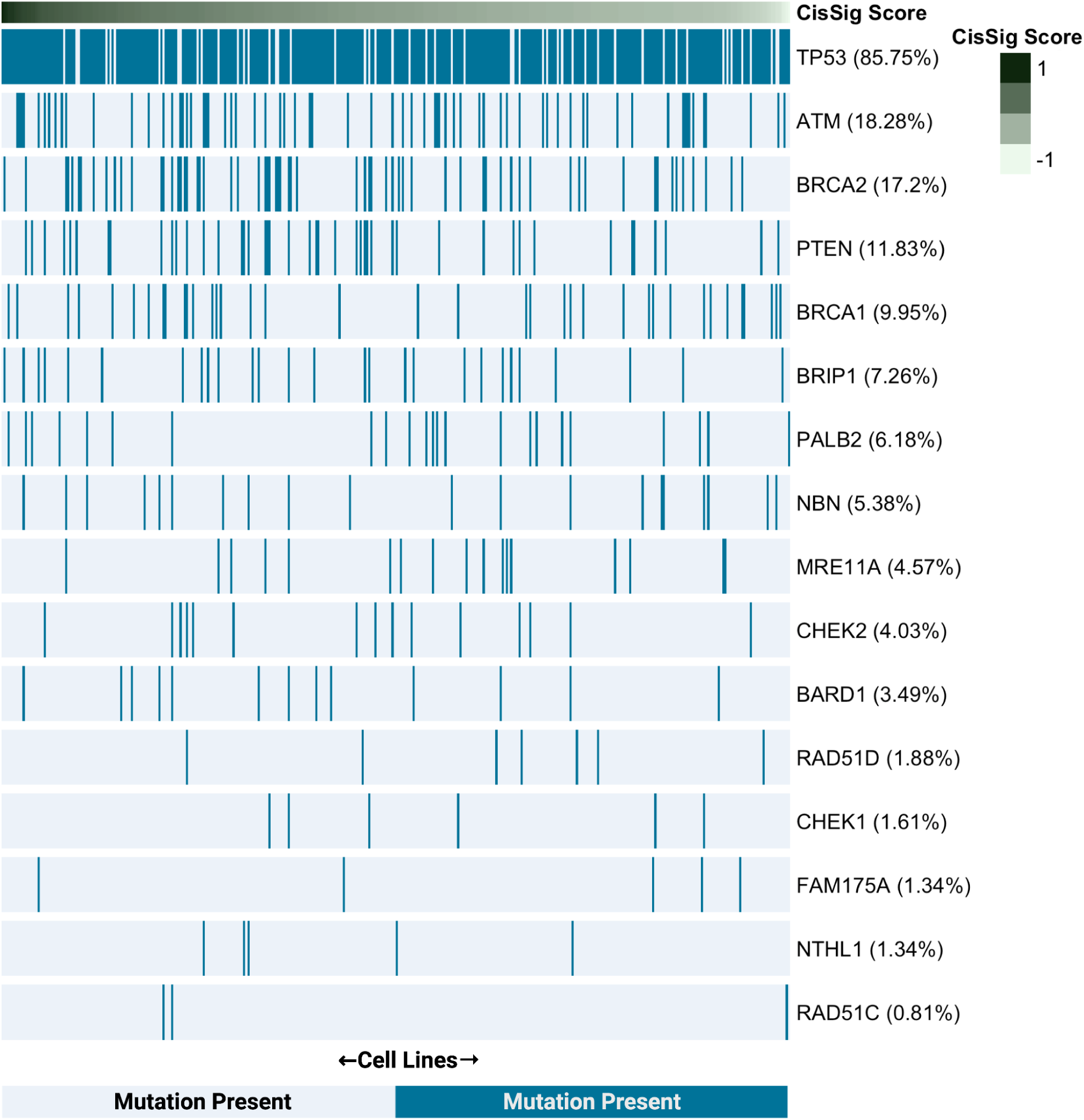
CisSig Score is not associated with mutation status in genes related to DNA damage response. A heatmap showing the mutation status for a variety of genes related to DNA damage response, in all epithelial based cell lines in the GDSC dataset. A cell line with the presence of any coding mutation is denoted as having a “mutation present.” Columns are ordered by CisSig Score, and rows are ordered by the frequency of a gene’s mutation within these cell lines. Mutation frequency is denoted in parentheses to the right of each gene name. After Bonferroni correction for multiple hypothesis testing, there is no statistical association (corrected p-value < 0.05 using chi-square test) found between cell lines in the top and bottom half of CisSig scores and the presence of a mutation in any of the genes.

**Figure S5.**
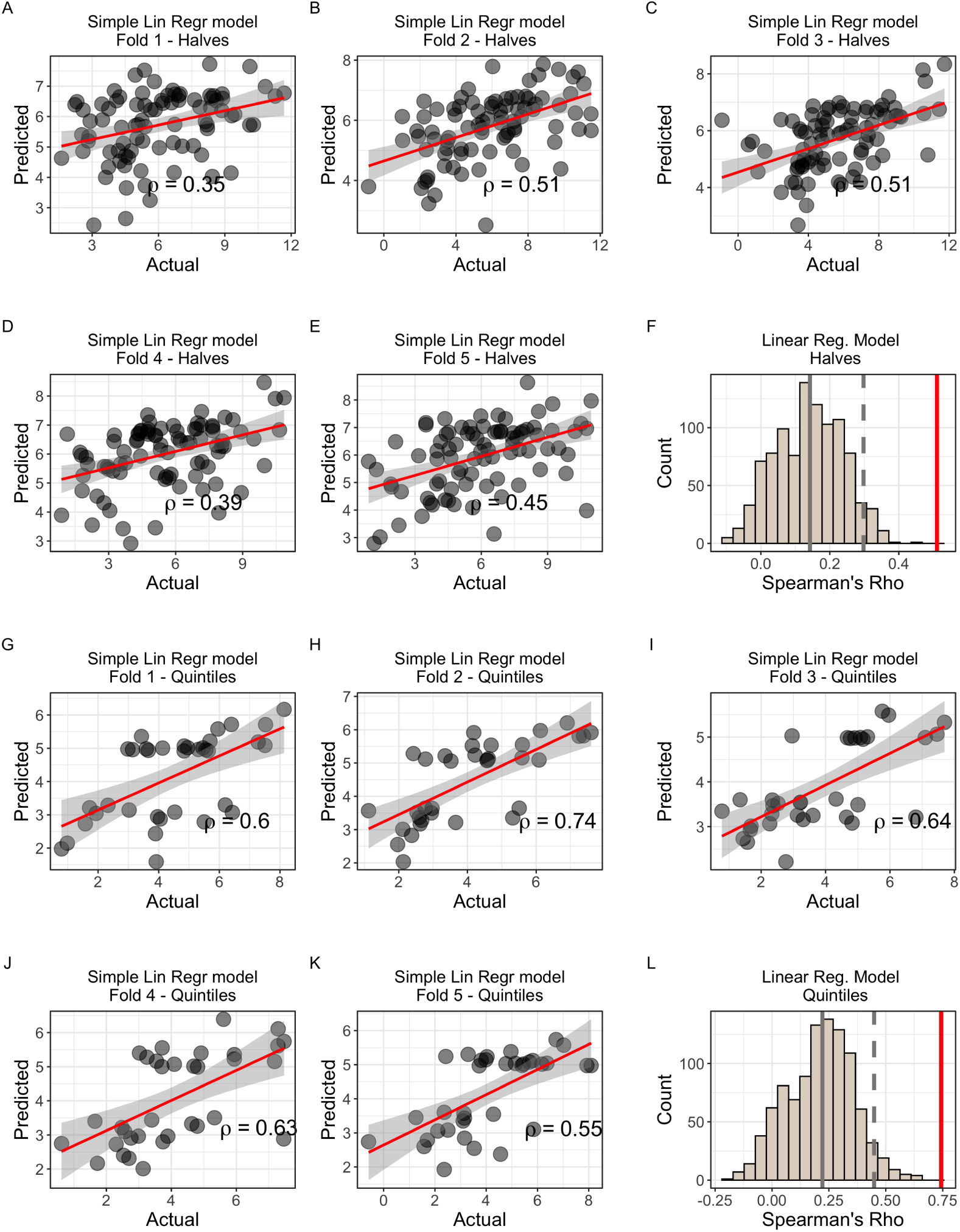
Modeling IC50 response using CisSig Score to predict IC50 in GDSC with simple linear regression. A-E. Predicted vs. Actual IC50 for validation sets of folds 1-5 for models built with all 429 cell lines. **F.** Null distribution of modeling metrics using 1000 random gene signatures with the same length as CisSig and the model described in **A-E**. CisSig’s performance (red solid line) is within the top 5% of the null distribution (cutoff at gray dashed line). Gray solid line represents median of null distribution. **G-K.** Predicted vs. Actual IC50 for validation sets of folds 1-5 for models built using cell lines in the top and bottom 20% of cisplatin IC50. **F.** Null distribution of modeling metrics using 1000 random gene signatures with the same length as CisSig and the model described in **G-K**. CisSig’s performance (red solid line) is compared to the 95% confidence interval (gray dashed line) of the null distribution. Gray solid line represents median of null distribution.

**Figure S6.**
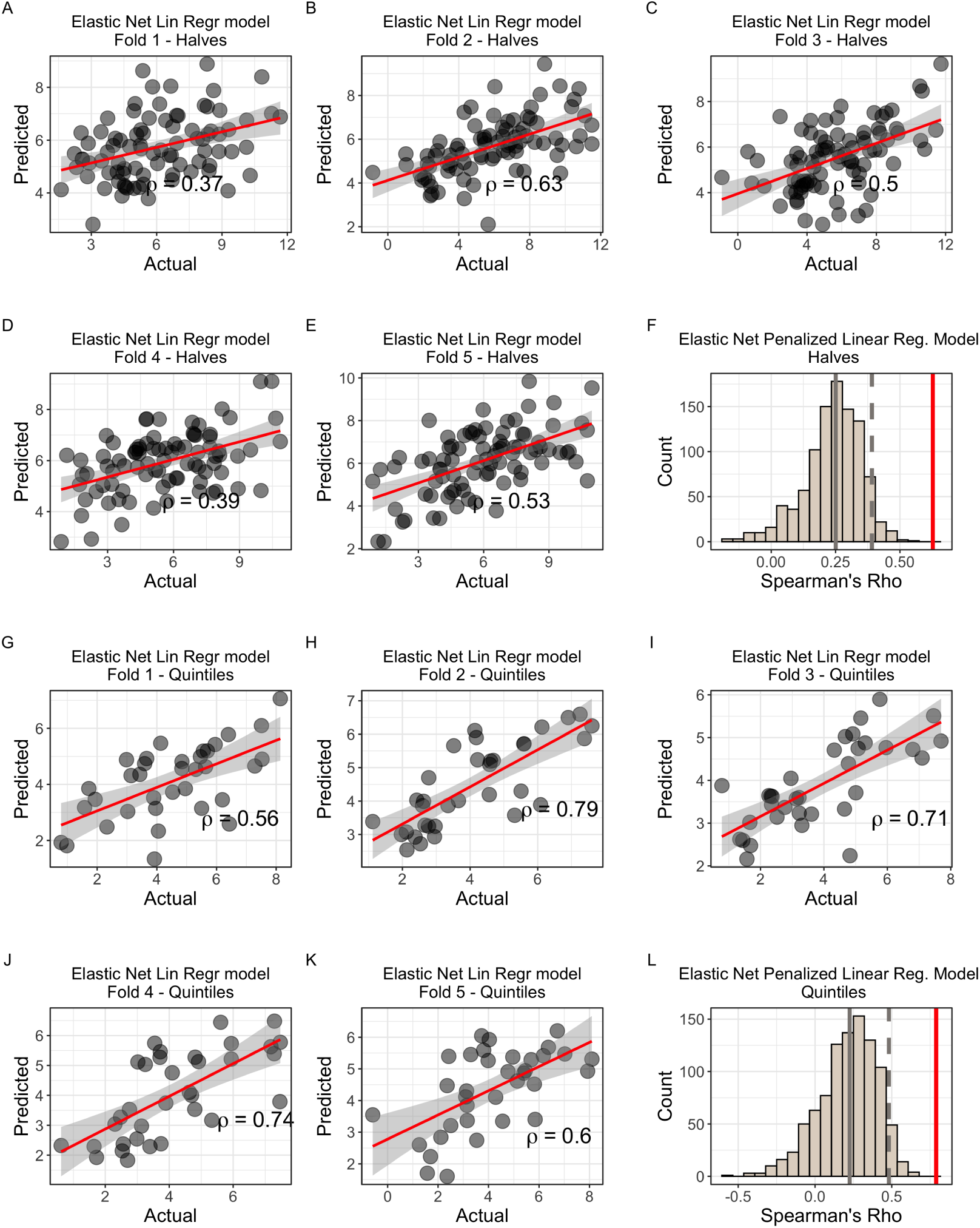
Modeling IC50 response using individual CisSig genes to predict IC50 in GDSC with elastic net penalized linear regression. A-E. Predicted vs. Actual IC50 for validation sets of folds 1-5 for models built with all 429 cell lines. F. Null distribution of modeling metrics using 1000 random gene signatures with the same length as CisSig and the model described in A-E. CisSig’s performance (red solid line) is within the top 5% of the null distribution (cutoff at gray dashed line). Gray solid line represents median of null distribution. G-K. Predicted vs. Actual IC50 for validation sets of folds 1-5 for models built using cell lines in the top and bottom 20% of cisplatin IC50. F. Null distribution of modeling metrics using 1000 random gene signatures with the same length as CisSig and the model described in G-K. CisSig’s performance (red solid line) is compared to the 95% confidence interval (gray dashed line) of the null distribution. Gray solid line represents median of null distribution.

**Figure S7.**
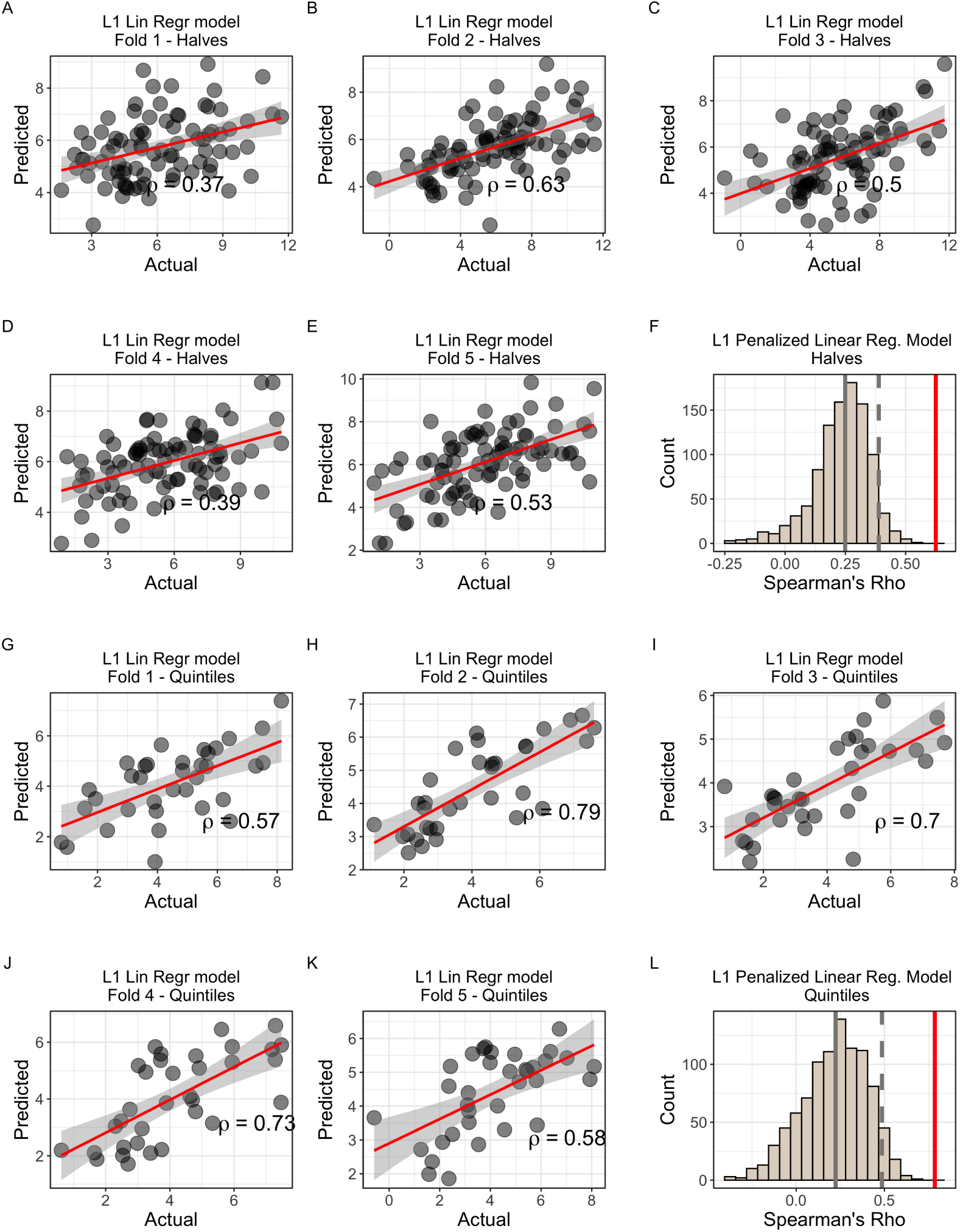
Modeling IC50 response using individual CisSig genes to predict IC50 in GDSC with L1 penalized linear regression. A-E. Predicted vs. Actual IC50 for validation sets of folds 1-5 for models built with all 429 cell lines. **F.** Null distribution of modeling metrics using 1000 random gene signatures with the same length as CisSig and the model described in **A-E**. CisSig’s performance (red solid line) is within the top 5% of the null distribution (cutoff at gray dashed line). Gray solid line represents median of null distribution. **G-K.** Predicted vs. Actual IC50 for validation sets of folds 1-5 for models built using cell lines in the top and bottom 20% of cisplatin IC50. **F.** Null distribution of modeling metrics using 1000 random gene signatures with the same length as CisSig and the model described in **G-K**. CisSig’s performance (red solid line) is compared to the 95% confidence interval (gray dashed line) of the null distribution. Gray solid line represents median of null distribution.

**Figure S8.**
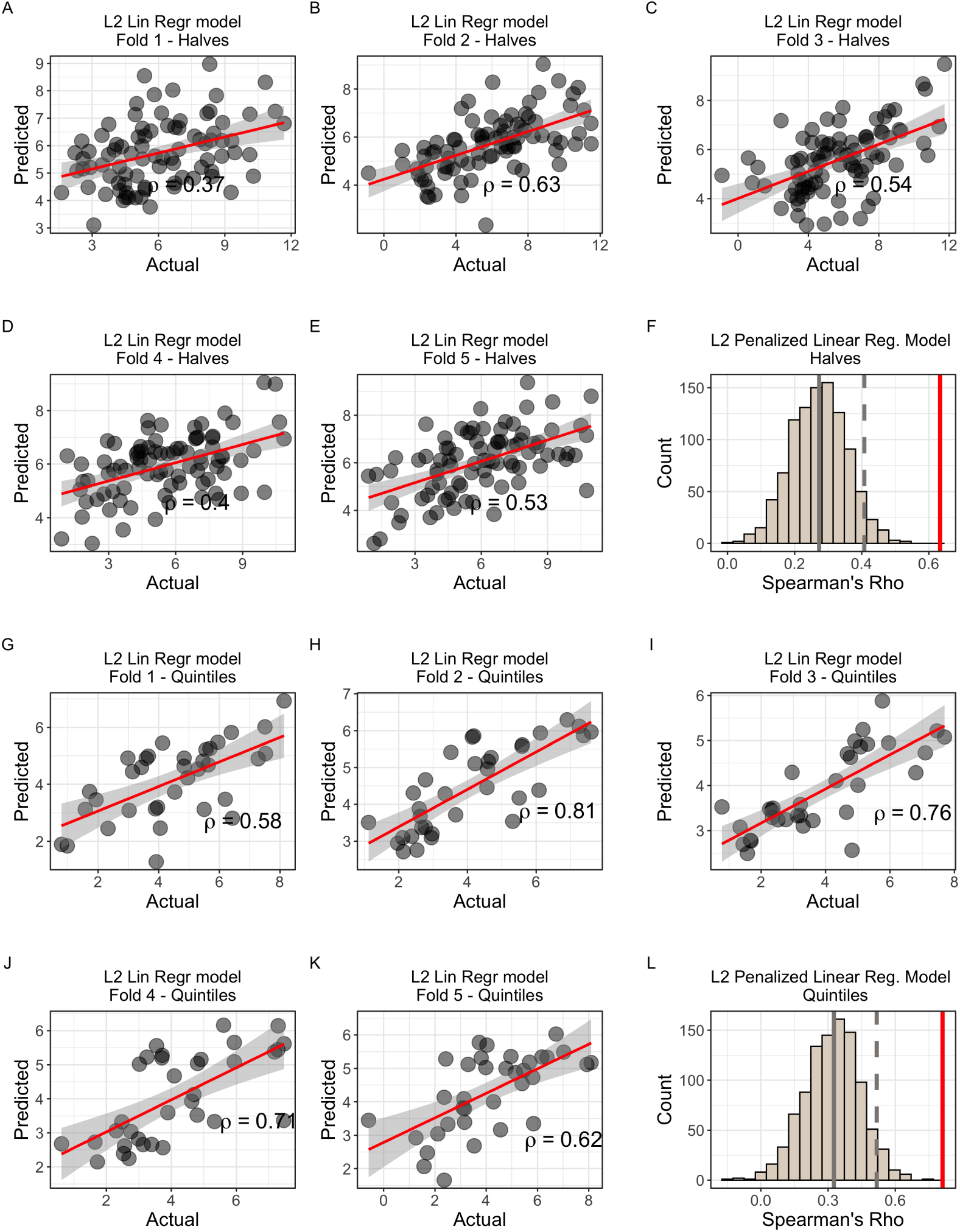
Modeling IC50 response using individual CisSig genes to predict IC50 in GDSC with L2 penalized linear regression. A-E. Predicted vs. Actual IC50 for validation sets of folds 1-5 for models built with all 429 cell lines. **F.** Null distribution of modeling metrics using 1000 random gene signatures with the same length as CisSig and the model described in **A-E**. CisSig’s performance (red solid line) is within the top 5% of the null distribution (cutoff at gray dashed line). Gray solid line represents median of null distribution. **G-K.** Predicted vs. Actual IC50 for validation sets of folds 1-5 for models built using cell lines in the top and bottom 20% of cisplatin IC50. **F.** Null distribution of modeling metrics using 1000 random gene signatures with the same length as CisSig and the model described in **G-K**. CisSig’s performance (red solid line) is compared to the 95% confidence interval (gray dashed line) of the null distribution. Gray solid line represents median of null distribution.

**Figure S9.**
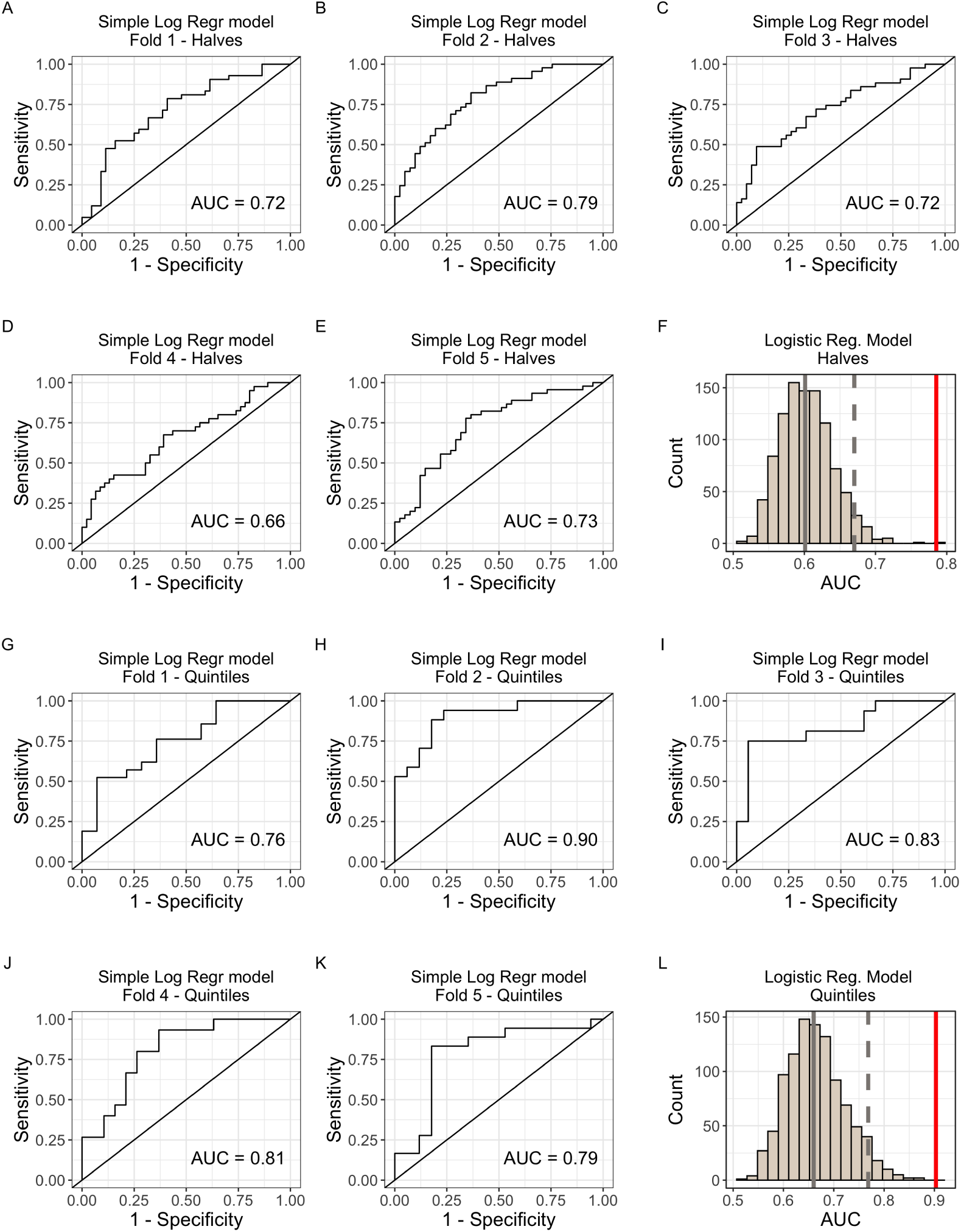
Modeling IC50 response using CisSig score to predict IC50 class in GDSC with simple logistic regression. A-E. Predicted vs. Actual IC50 for validation sets of folds 1-5 for models built with all 429 cell lines. **F.** Null distribution of modeling metrics using 1000 random gene signatures with the same length as CisSig and the model described in **A-E**. CisSig’s performance (red solid line) is within the top 5% of the null distribution (cutoff at gray dashed line). Gray solid line represents median of null distribution. **G-K.** Predicted vs. Actual IC50 for validation sets of folds 1-5 for models built using cell lines in the top and bottom 20% of cisplatin IC50. **F.** Null distribution of modeling metrics using 1000 random gene signatures with the same length as CisSig and the model described in **G-K**. CisSig’s performance (red solid line) is compared to the 95% confidence interval (gray dashed line) of the null distribution. Gray solid line represents median of null distribution.

**Figure S10.**
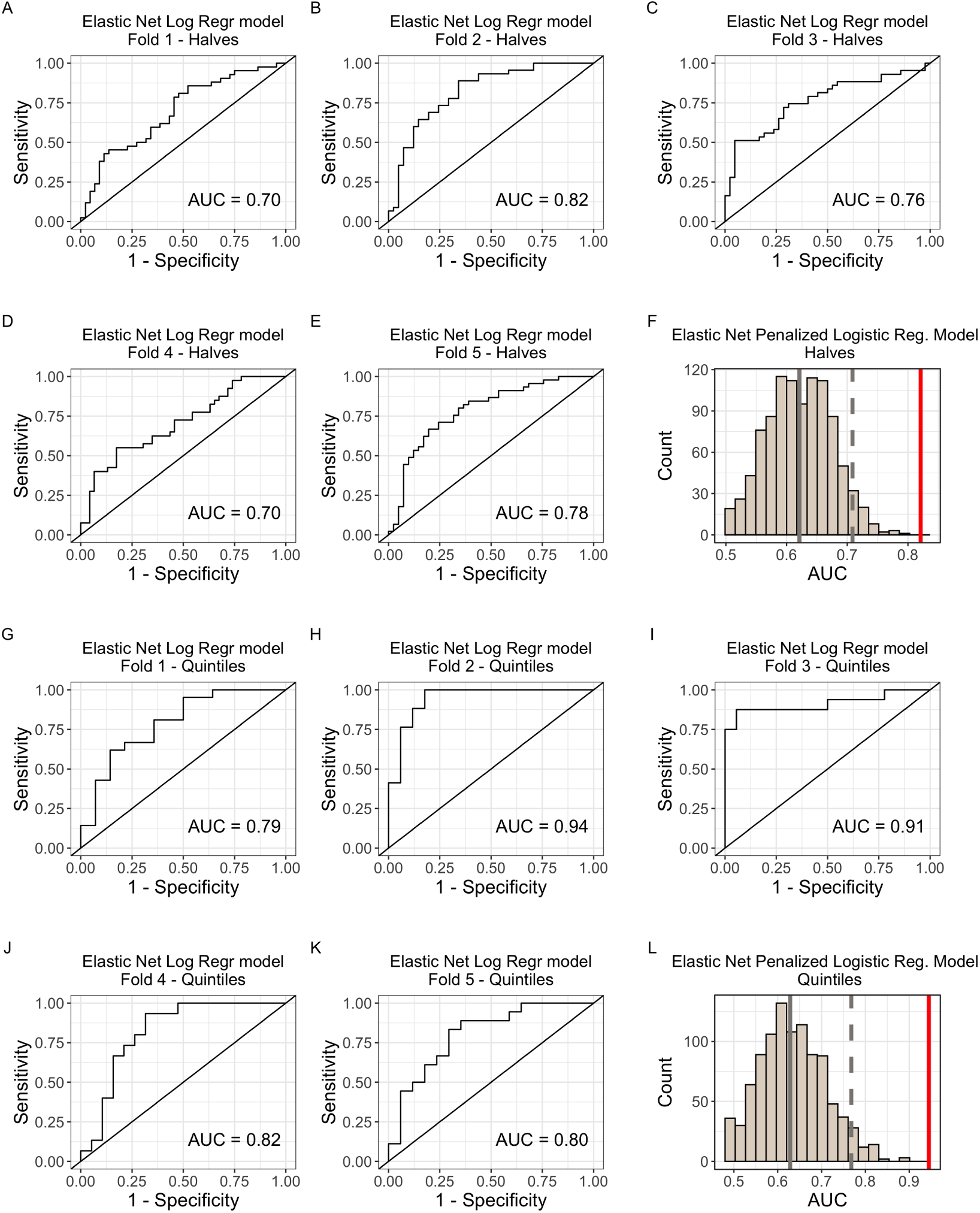
Modeling IC50 response using individual CisSig genes to predict IC50 class in GDSC with elastic net penalized logistic regression. A-E. Predicted vs. Actual IC50 for validation sets of folds 1-5 for models built with all 429 cell lines. F. Null distribution of modeling metrics using 1000 random gene signatures with the same length as CisSig and the model described in A-E. CisSig’s performance (red solid line) is within the top 5% of the null distribution (cutoff at gray dashed line). Gray solid line represents median of null distribution. G-K. Predicted vs. Actual IC50 for validation sets of folds 1-5 for models built using cell lines in the top and bottom 20% of cisplatin IC50. F. Null distribution of modeling metrics using 1000 random gene signatures with the same length as CisSig and the model described in G-K. CisSig’s performance (red solid line) is compared to the 95% confidence interval (gray dashed line) of the null distribution. Gray solid line represents median of null distribution.

**Figure S11.**
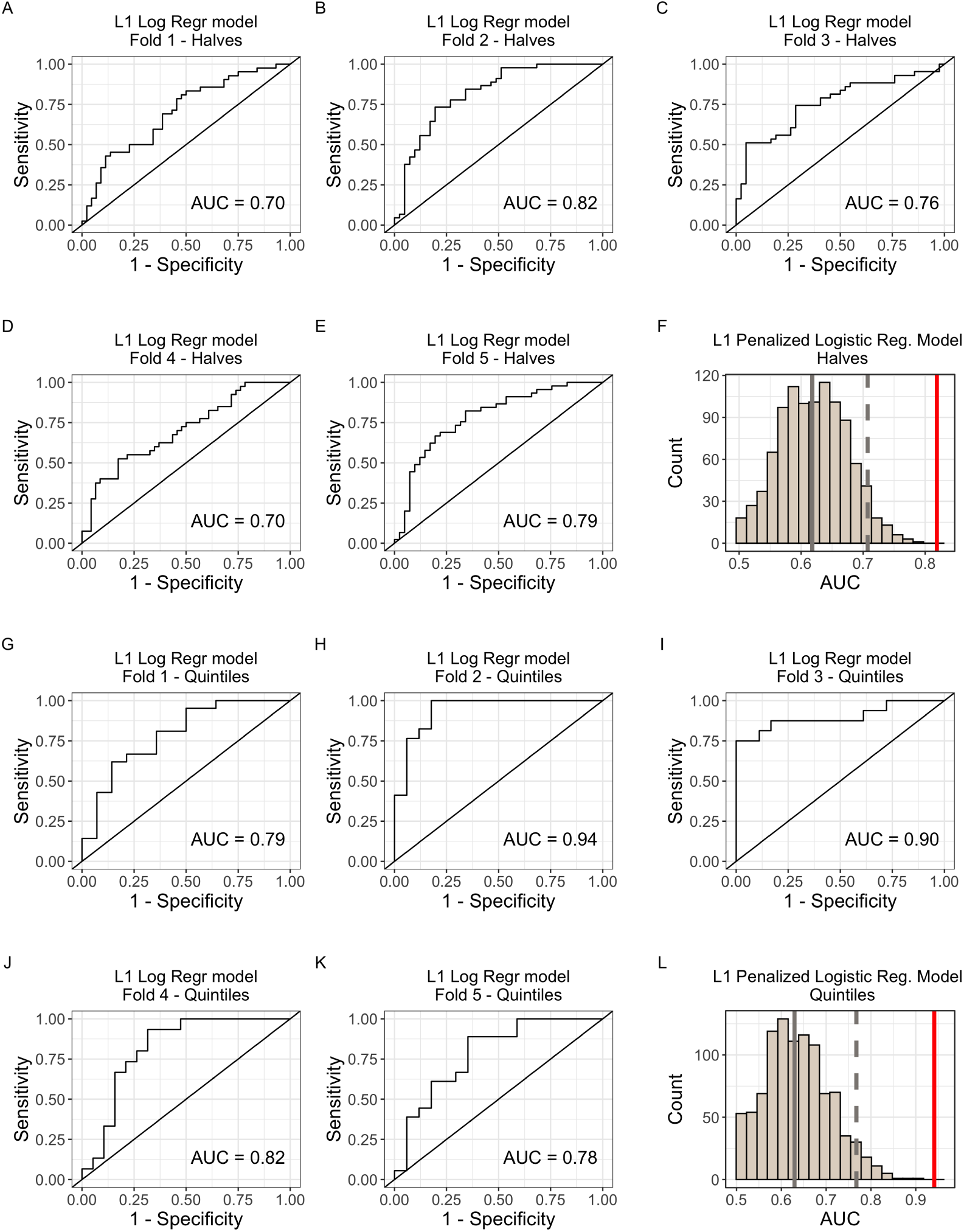
Modeling IC50 response using individual CisSig genes to predict IC50 class in GDSC with L1 penalized logistic regression. A-E. Predicted vs. Actual IC50 for validation sets of folds 1-5 for models built with all 429 cell lines. F. Null distribution of modeling metrics using 1000 random gene signatures with the same length as CisSig and the model described in A-E. CisSig’s performance (red solid line) is within the top 5% of the null distribution (cutoff at gray dashed line). Gray solid line represents median of null distribution. G-K. Predicted vs. Actual IC50 for validation sets of folds 1-5 for models built using cell lines in the top and bottom 20% of cisplatin IC50. F. Null distribution of modeling metrics using 1000 random gene signatures with the same length as CisSig and the model described in G-K. CisSig’s performance (red solid line) is compared to the 95% confidence interval (gray dashed line) of the null distribution. Gray solid line represents median of null distribution.

**Figure S12.**
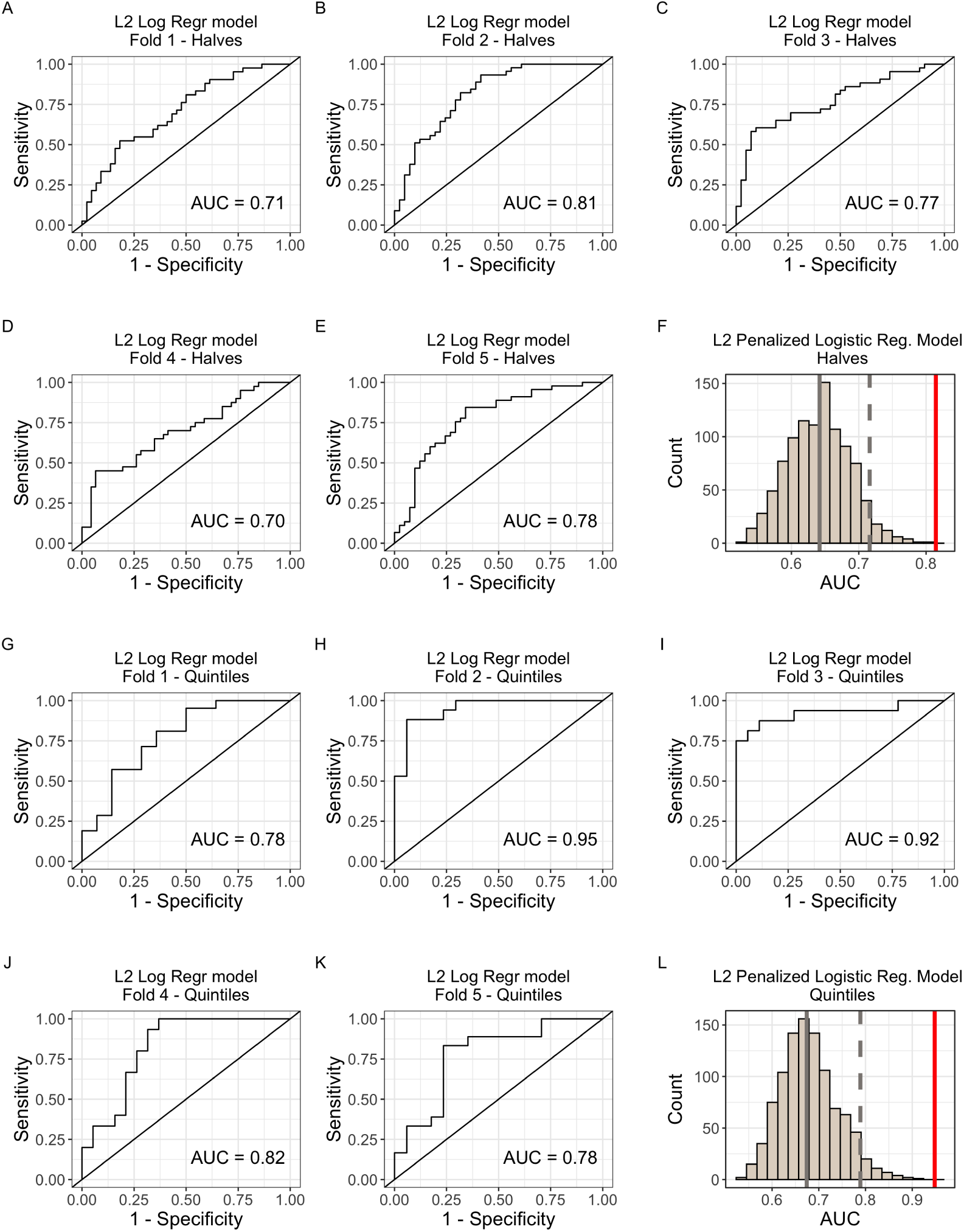
Modeling IC50 response using individual CisSig genes to predict IC50 class in GDSC with L2 penalized logistic regression. A-E. Predicted vs. Actual IC50 for validation sets of folds 1-5 for models built with all 429 cell lines. F. Null distribution of modeling metrics using 1000 random gene signatures with the same length as CisSig and the model described in A-E. CisSig’s performance (red solid line) is within the top 5% of the null distribution (cutoff at gray dashed line). Gray solid line represents median of null distribution. G-K. Predicted vs. Actual IC50 for validation sets of folds 1-5 for models built using cell lines in the top and bottom 20% of cisplatin IC50. F. Null distribution of modeling metrics using 1000 random gene signatures with the same length as CisSig and the model described in G-K. CisSig’s performance (red solid line) is compared to the 95% confidence interval (gray dashed line) of the null distribution. Gray solid line represents median of null distribution.

**Figure S13.**
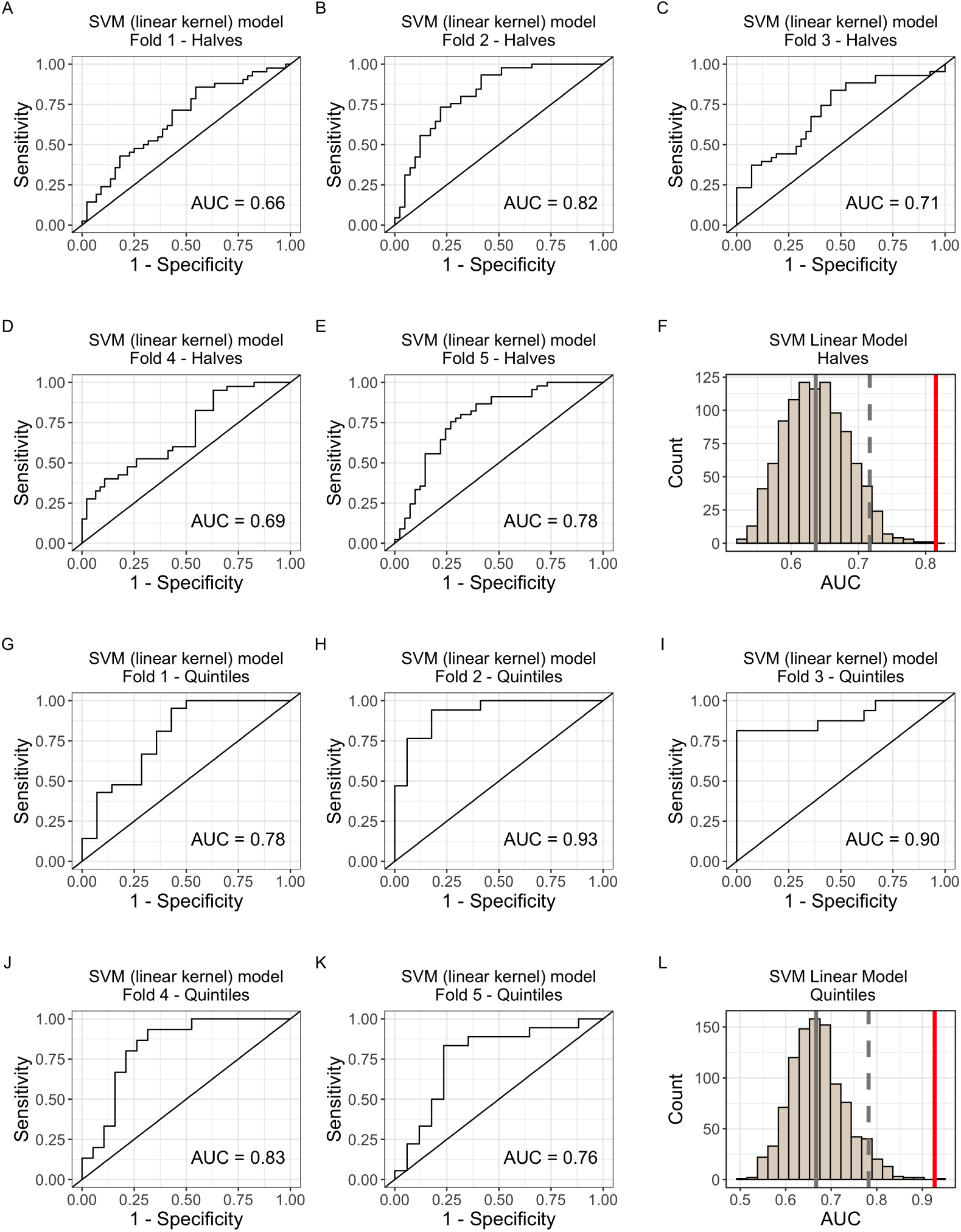
Modeling IC50 response using individual CisSig genes to predict IC50 class in GDSC with support vector machine modeling (linear kernel). A-E. Predicted vs. Actual IC50 for validation sets of folds 1-5 for models built with all 429 cell lines. F. Null distribution of modeling metrics using 1000 random gene signatures with the same length as CisSig and the model described in A-E. CisSig’s performance (red solid line) is within the top 5% of the null distribution (cutoff at gray dashed line). Gray solid line represents median of null distribution. G-K. Predicted vs. Actual IC50 for validation sets of folds 1-5 for models built using cell lines in the top and bottom 20% of cisplatin IC50. F. Null distribution of modeling metrics using 1000 random gene signatures with the same length as CisSig and the model described in G-K. CisSig’s performance (red solid line) is compared to the 95% confidence interval (gray dashed line) of the null distribution. Gray solid line represents median of null distribution.

**Figure S14.**
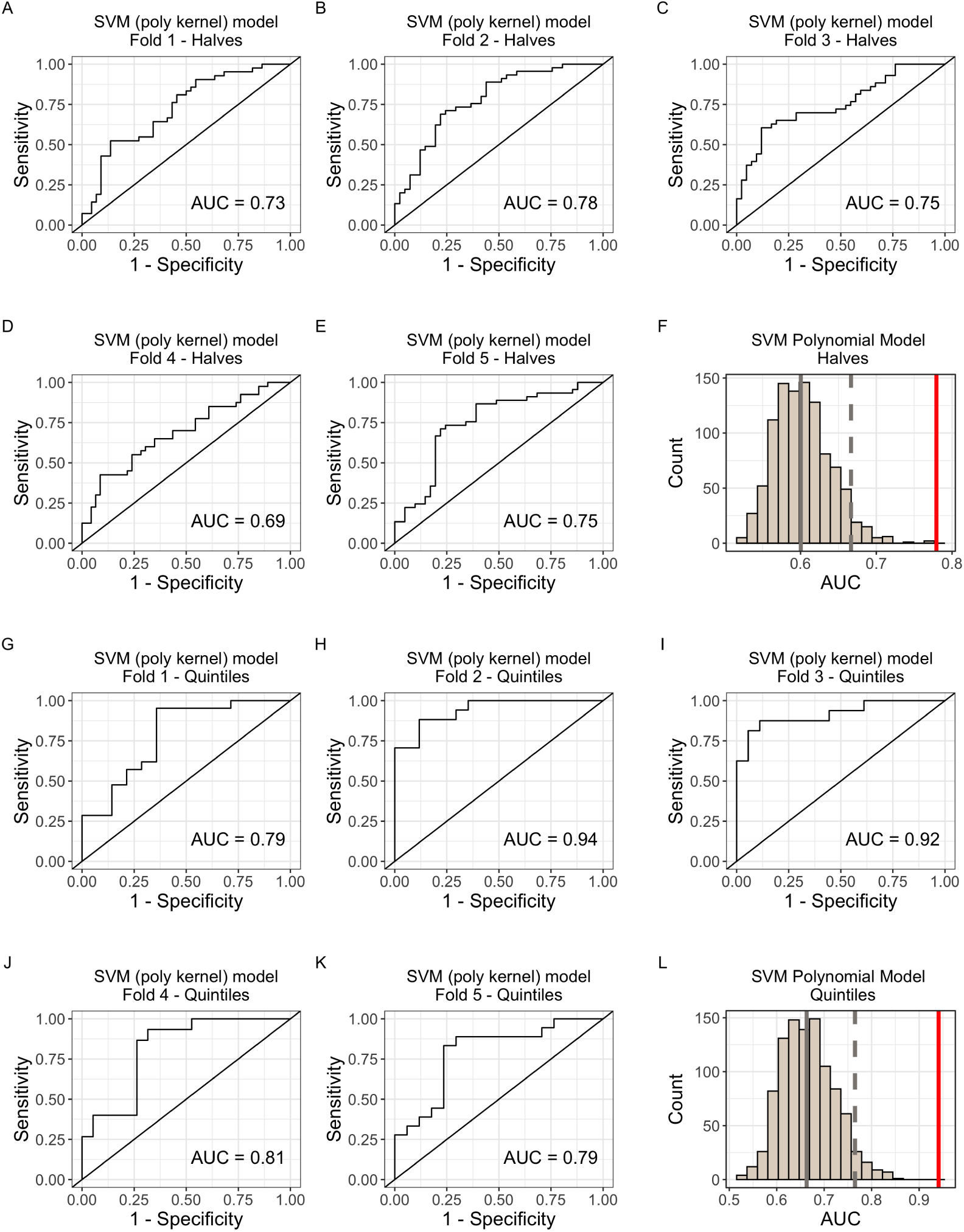
Modeling IC50 response using individual CisSig genes to predict IC50 class in GDSC with support vector machine modeling (polynomial kernel). A-E. Predicted vs. Actual IC50 for validation sets of folds 1-5 for models built with all 429 cell lines. F. Null distribution of modeling metrics using 1000 random gene signatures with the same length as CisSig and the model described in A-E. CisSig’s performance (red solid line) is within the top 5% of the null distribution (cutoff at gray dashed line). Gray solid line represents median of null distribution. G-K. Predicted vs. Actual IC50 for validation sets of folds 1-5 for models built using cell lines in the top and bottom 20% of cisplatin IC50. F. Null distribution of modeling metrics using 1000 random gene signatures with the same length as CisSig and the model described in G-K. CisSig’s performance (red solid line) is compared to the 95% confidence interval (gray dashed line) of the null distribution. Gray solid line represents median of null distribution.

**Figure S15.**
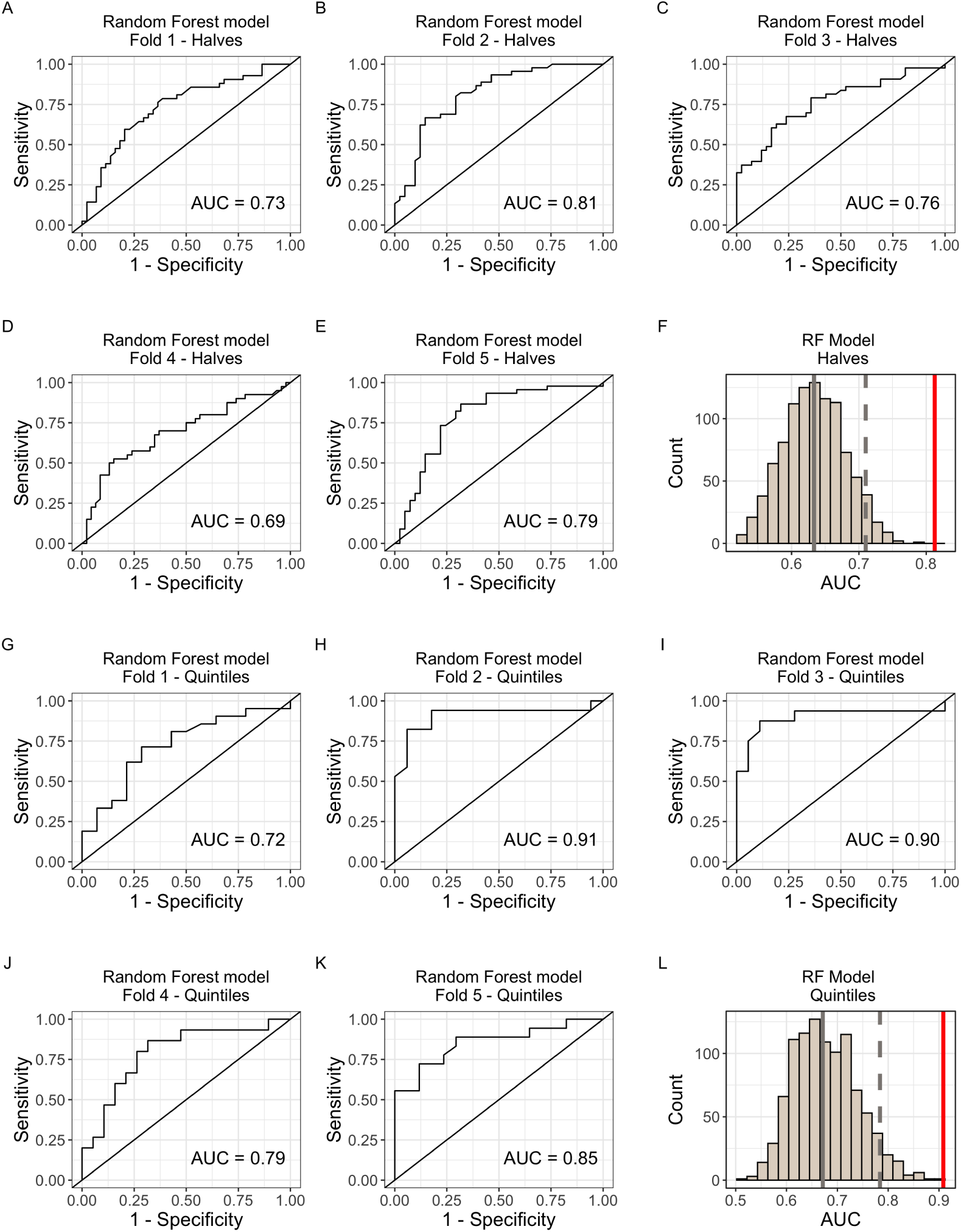
Modeling IC50 response using individual CisSig genes to predict IC50 class in GDSC with random forest modeling. A-E. Predicted vs. Actual IC50 for validation sets of folds 1-5 for models built with all 429 cell lines. **F.** Null distribution of modeling metrics using 1000 random gene signatures with the same length as CisSig and the model described in **A-E**. CisSig’s performance (red solid line) is within the top 5% of the null distribution (cutoff at gray dashed line). Gray solid line represents median of null distribution. **G-K.** Predicted vs. Actual IC50 for validation sets of folds 1-5 for models built using cell lines in the top and bottom 20% of cisplatin IC50. **F.** Null distribution of modeling metrics using 1000 random gene signatures with the same length as CisSig and the model described in **G-K**. CisSig’s performance (red solid line) is compared to the 95% confidence interval (gray dashed line) of the null distribution. Gray solid line represents median of null distribution.

**Figure S16.**
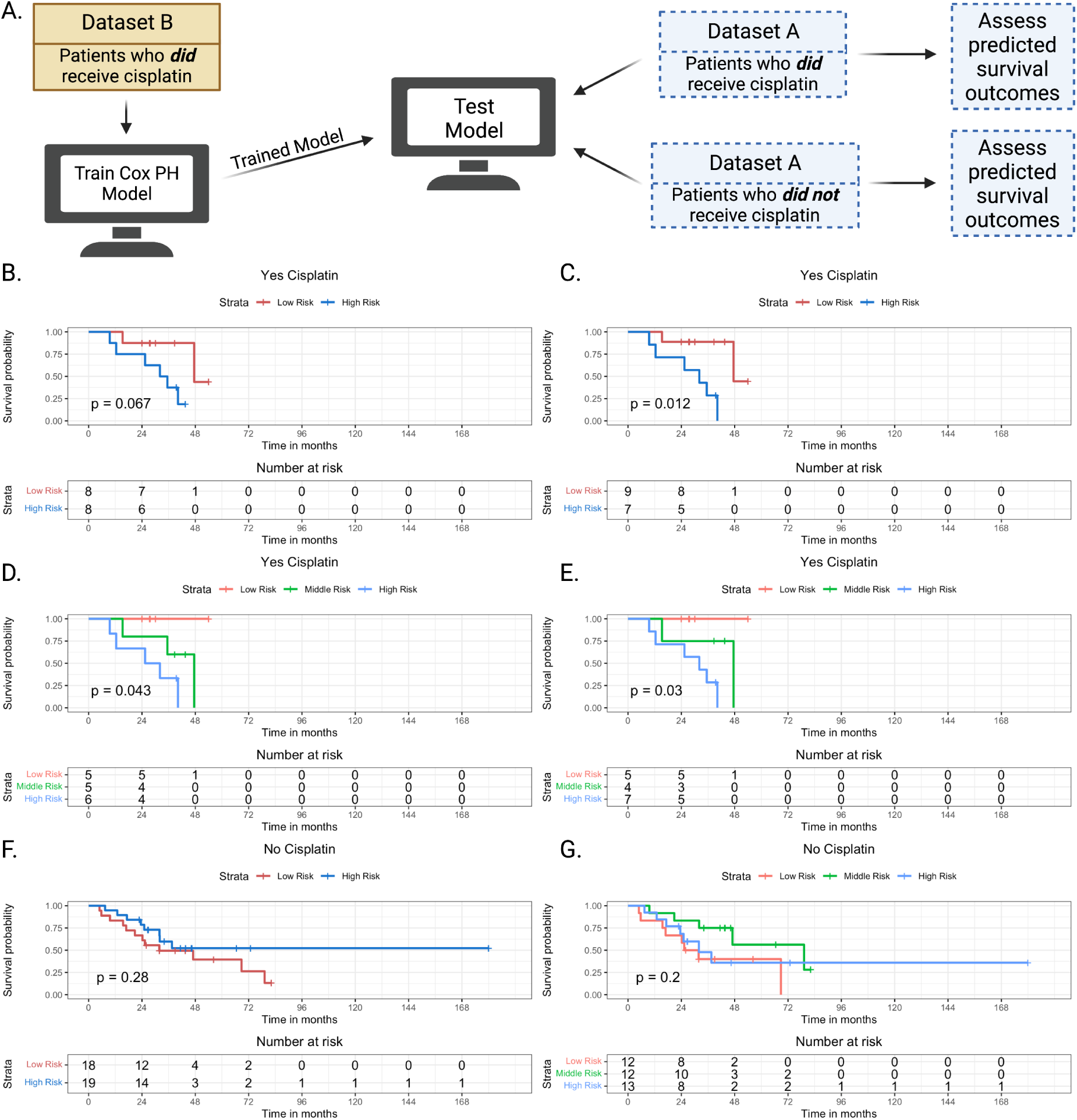
CisSig-trained model is predictive in patients who have received cisplatin, but lacks signal in patients who have not received cisplatin. A. Schematic description of model training and testing, where model is trained using patients who did receive cisplatin-containing treatment from Dataset B. Testing of the trained model is done using patients from the Dataset A who did not receive cisplatin-containing treatment and patients from the Dataset A who did receive cisplatin-containing treatment. B. Test samples that did receive cisplatin-containing treatment are separated into groups of “high” and “low risk” based on the model’s predictions using a median cutoff. Kaplan-meier curves show a significant separation between the two groups. C. The same analysis shown in B, using an optimal cutpoint (determined by chi-square statistic) instead of median to separate the cohorts. D-E. The same analyses shown in B-C, separating the groups into “high”, “middle”, and “low risk” groups using tertiles and the optimal two cutpoints, respectively. F-G. The same analyses shown in B and D, using samples from Dataset A that did not receive cisplatin-containing treatment, demonstrating no significant separation between the two groups.

